# Association Between Anorexia Nervosa Severity and Social and Occupational Functioning across Adulthood

**DOI:** 10.64898/2026.11.05.26365199

**Authors:** Zi-Ping Zhang, Janne Tidselbak Larsen, Loa Clausen, Esben Agerbo, Cynthia M. Bulik, Liselotte Vogdrup Petersen, Zeynep Yilmaz

## Abstract

**Importance:** Severe anorexia nervosa (AN) carries substantial mortality risk, but its consequences associated with social and occupational functioning remain poorly characterized, limiting the development of targeted interventions to improve functioning, prevent and reduce long-term social disadvantage.

**Objective:** To determine whether AN severity is associated with educational attainment, employment, income, and cohabitation status.

**Design:** Cohort study following social and occupational outcomes across adulthood until age 55.

**Setting:** Population-based.

**Participants:** All individuals born in Denmark from 1963 to 1994, alive and residing in Denmark at age 30. The AN group had a first AN diagnosis between ages 10 and 25 years, excluding those who died or emigrated within 5 years. The comparison group had no AN diagnosis.

**Exposure:** Illness severity of AN, measured by the AN Register-based Severity Index 5 years after the first AN diagnosis.

**Main Outcomes and Measures:** Educational attainment was assessed at age 25 (upper secondary education) and at age 30/35 (highest completed education). Employment status, income deficiency, and cohabitation status were assessed annually from age 30 to 55.

**Results:** Among 2,442,916 individuals, 9,465 were diagnosed with AN, of whom 1,996 (21.1%) were classified as severe AN cases. At age 25, individuals with severe AN had increased odds of non-completion of upper-secondary education compared to the general population (odds ratio [OR]=3.07; 95% confidence interval [CI]: 2.72-3.46), and those with less severe AN (OR=1.39; 95% CI: 1.29-1.50). At age 30, severe AN was associated with higher odds of unemployment (OR=3.42; 95% CI: 3.03-3.86), income deficiency (OR=3.84; 95% CI: 3.42-4.31), and living alone (OR=2.43; 95% CI: 2.17-2.71) compared to the less severe AN group. The risk of unemployment and income deficiency diverged across time: individuals with less severe AN gradually approached the general population, whereas those with severe AN diverged further over time. Within the AN cohort, severity differences persisted after adjustment for active AN diagnosis, with minimal attenuation.

**Conclusions and Relevance:** Greater lifetime AN severity was associated with significantly poorer long-term social and occupational functioning outcomes, and these functional difficulties may persist beyond periods of active illness. This vulnerable group needs substantial support in these areas throughout the course of illness.

**Key points:** *Question:* Is anorexia nervosa (AN) severity associated with long-term social and occupational functioning outcomes?

*Findings:* In this population-based cohort study, greater AN severity was associated with poorer long-term functioning, including education, occupation, and cohabitation status, across adulthood. AN severity showed graded, dose-response-like relationships with those outcomes, and the gradient persisted regardless of whether individuals had active eating disorder diagnoses.

*Meaning:* Individuals with severe AN show substantial social and occupational impairment, reflected in shorter education, weaker labour-market attachment, lower income, and greater likelihood of living alone, compared with the general population, with these disparities becoming more pronounced over time.

## Introduction

Anorexia nervosa (AN) is a serious psychiatric disorder associated with a high likelihood of chronicity, multimorbidity, and premature mortality.^[1, 2]^ Beyond its clinical impact, AN disrupts key developmental stages and impairs psychosocial functioning, often generating lasting socioeconomic and functional impairment.^[3–5]^ While Global Burden of Disease data indicate that years lived with disability (YLD) for many mental disorders are stable or declining, YLD attributable to AN has increased over the past three decades.^[6, 7]^ Most prior research has examined educational attainment and socioeconomic status as risk factors preceding AN onset,^[8, 9]^ whereas few studies have examined them as consequences of AN. Register-based studies consistently report adverse outcomes following AN diagnosis, though findings vary in magnitude. A Swedish cohort study found that 58% of individuals with AN were employed and 21% received disability pension or other sickness benefits 9 to 14 years after hospital admission.^[4]^ A Finnish 30-year follow-up study similarly reported that 22% of individuals with AN received disability pension, alongside reduced employment days and lower cumulative income relative to population controls.^[3]^ Educational trajectories may also be altered: females with AN being overrepresented at both the lowest and highest levels of attainment compared to matched controls.^[10]^ A separate community-based cohort found that more women with AN experienced delayed education, remaining enrolled in higher education into their early thirties.^[11]^

Past studies have relied on AN diagnosis alone without accounting for heterogeneity in symptom severity, chronicity, and treatment response. Nonetheless, some evidence suggests the severity itself matters: higher eating disorders (EDs) symptom levels were associated with worse occupational functioning in a female sample.^[12]^ Consistent with this, researchers observed that adults with severe and enduring AN had poor baseline work and social adjustment, followed by limited improvement in the short term.^[13]^ Despite this evidence, it remains unclear whether these associations reflect a severity gradient or a disproportionate burden specific to the most severely affected, and/or whether a potential gradient persists after remission. Understanding this relationship could potentially identify patients at greatest risk of long-term disadvantage, with relevance for service provision.

Understanding the impact of severe AN has been limited by the lack of a validated severity measurement in epidemiological research.^[14–16]^ To remedy this, we derived a continuous composite score using information from the Danish national registers. The Anorexia Nervosa Register-based Severity Index (AN-RSI) enables quantification of illness severity in population-based register studies^[17]^ and significantly predicts mortality ^[17]^, supporting its utility as an indicator of overall cumulative illness burden. The features it may reflect, including prolonged undernutrition, frequent comorbidity, and repeated treatment exposure, also plausibly drive long-term socioeconomic disadvantage,^[18–20]^ making AN-RSI well-suited to examining how illness severity shapes long-term socio-occupational functioning in AN.

The primary aim of our study was to examine the severity-stratified relationship with long-term social and occupational outcomes in AN. We hypothesized that more severe AN would be associated with greater cumulative impairment in educational, occupational, and social functioning. Specifically, we: (1) quantified the burden of AN across severity levels by comparing multiple outcomes including educational attainment, employment status, income level, and cohabitation status against the general population; and (2) evaluated the association between illness severity and functional outcomes among individuals with AN.

## Methods

### Study design and data

This register-based cohort study used individual-level data from Danish national registers, linked via the unique personal identification numbers assigned to all Danish residents through the Danish Civil Registration System.^[21]^ All live-born individuals in Denmark between 1963 and 1994 were followed from age 25 until age 55 or through December 31, 2024. Individuals with a first AN diagnosis (International Statistical Classification of Diseases (ICD), including ICD-10: F50.0-F50.1 and ICD-8: 306.50) were identified from the Danish National Patient Register and the Danish Psychiatric Central Research Register.^[22, 23]^ Given the timing of education completion and career entry, we excluded individuals older than 25 years at first AN diagnosis. To ensure comparable exposure time for the AN-RSI calculation (eMethods), we also excluded individuals younger than 10 years. Individuals who died or emigrated within 5 years of first diagnosis were excluded, since AN-RSI requires 5 years of complete follow-up. Educational attainment was obtained from the Danish Education Registers,^[24]^ and employment and income data were obtained from the Employment Classification Module and the Integrated Database for Labor-market Research.^[25]^ Cohabitation status was derived from the Danish Population Statistics Register.^[26]^ Full register descriptions appear in the eMethods. This study followed the STROBE reporting guideline.^[27]^

The AN-RSI incorporates age of onset, illness duration, treatment length, and healthcare utilization. Illness severity was quantified using the AN-RSI five years after the first AN diagnosis following the weighting procedure described elsewhere^[17]^ and detailed in the eMethods. Individuals with AN were stratified in two ways. In the binary classification, cases scoring in the top quintile of the AN-RSI were classified as severe AN and the remainder as less severe AN, consistent with the proportion of individuals who experience a severe illness course reported in previous studies.^[28]^ The severity scores were further divided into quintiles.

We examined four socio-occupational domains. In Denmark, upper-secondary education is typically completed by age 20, and most people have achieved their highest educational attainment by age 30.^[29]^ To account for minor delays, we measured: (1) completion of upper-secondary education at age 25 (binary); and (2) highest educational attainment at age 35 (four levels, ordinal). For labour-market and social outcomes, we measured employment status (unemployment or employment), income deficiency (lowest quintile of annual wages, relative to the age-, sex-, and calendar year-matched general population), and cohabitation status (living alone or not) as trajectories from ages 30 to 55. Detailed outcome definitions and rationales are provided in eMethods.

### Statistical analyses

To quantify the absolute burden in relation to functioning outcomes, we compared the AN severity group against the reference group (all individuals from the general population without a diagnosis of AN). We examined the associations between education outcomes and illness severity at ages 25 and 30 with logistic regression for binary outcomes and multinomial logistic regression for categorical outcomes. Trajectories of employment status, income level, and cohabitation status were analyzed by fitting separate regressions for every year between 30 and 55. To evaluate whether long-term socio-occupational impairments were driven by ongoing, active diagnoses, we conducted within-AN analyses using three nested models: a crude model adjusted for baseline covariates; a model additionally adjusted for active AN, defined as an inpatient or outpatient contact with an AN diagnosis recorded during the calendar year in which the outcome was measured; and an alternative model adjusted for active ED, defined analogously using any ED diagnosis. The latter adjustment was performed as a sensitivity analysis to address the potential diagnostic crossover. To examine whether illness severity showed a graded association with functional outcomes, additional analyses were performed using the AN-RSI quintiles as exposure.

All models were adjusted for sex, birth year, parental psychiatric history, and socioeconomic position, which were selected a priori based on previous literature and confounding (eMethods). Complete-case analysis was used after excluding missing covariate data. Statistical tests were two-sided with Benjamini-Hochberg false discovery rate correction per outcome, performed in R version 4.4.3.^[30]^

## Results

In total, 2,442,916 individuals were included in our study, of whom 9,465 were diagnosed with AN between ages 10 and 25; 1,996 (21.1%) met the AN-RSI threshold for severe AN (eTable 1-2). The AN cohort was predominantly female (93.8%), with a mean age at first diagnosis of 17.9 years (standard deviation [SD]=3.5). Parents of the AN cohort had higher income, higher secondary education completion, and more frequent psychiatric history compared to the general population (Table 1).

**Table 1.** Baseline characteristics of study participants.

| **Characteristic** | **AN, total**<br>n = 9,465 | **AN, severe**<br>n = 1,996 | **AN, less severe**<br>n = 7,469 | **General population**<sup>a</sup><br>n = 2,433,451 |
|---|---|---|---|---|
| Female, No. (%) | 8,874 (93.8) | 1,895 (94.9) | 6,979 (93.4) | 1,186,388 (48.8) |
| Alive and resident at end of follow-up, No. (%) | 9,208 (97.3) | 1,936 (97.0) | 7,272 (97.4) | 2,352,750 (96.7) |
| Age at first diagnosis, mean (SD) | 17.9 (3.5) | 17.5 (3.6) | 18.0 (3.5) | — |
| AN-RSI score, mean (SD) | 5.0 (7.6) | 15.4 (10.9) | 2.3 (2.1) | — |
| Parental socioeconomic status<sup>b</sup> |  |  |  |  |
| Income deficiency, No. (%) | 2,626 (27.7) | 490 (24.5) | 2,136 (28.6) | 804,598 (33.1) |
| Unemployment, No. (%) | 2,679 (28.3) | 529 (26.5) | 2,150 (28.8) | 705,699 (29.0) |
| No secondary education, No. (%) | 3,093 (32.7) | 604 (30.3) | 2,489 (33.3) | 1,117,796 (45.9) |
| Parental psychiatric history, No. (%) | 2,820 (29.8) | 609 (30.5) | 2,211 (29.6) | 528,206 (21.7) |

AN: anorexia nervosa; AN-RSI: AN Register-based Severity Index; IQR: interquartile range; SD: standard deviation; a. general population without an AN diagnosis; b. parental socioeconomic status is measured at age 25 of the proband (see eMethods for details).

### Educational outcomes

By age 25, 527 individuals (35.9%) with severe AN had not completed upper-secondary education compared with 1,198 (23.0%) with less severe AN and 557,760 (22.9%) in the general population. (Table 2a) Both AN groups had a higher likelihood of not completing upper-secondary education than the general population, but the risk was twice as high in the severe group compared to the less severe group (odds ratio [OR]=3.07; 95% CI:2.72-3.46; OR =1.39; 95% CI:1.29-1.50, respectively).

**Table 2a.** Odds ratio of not having upper-secondary education by anorexia nervosa severity at age 25.

| Education | Group | No. (%) | Odds ratio<br>(95% CI) | P value |
|---|---|---|---|---|
| Not having upper-secondary education | General population | 557,760 (22.9%) | 1 [Reference] |  |
|  | Less severe AN | 1,198 (23.0%) | 1.39 (1.29-1.50) | <.001 |
|  | Severe AN | 527 (35.9%) | 3.07 (2.72-3.46) | <.001 |

AN: anorexia nervosa; CI: confidence interval.

**Table 2b.** Odds ratio of highest educational level by anorexia nervosa severity at age 35.

| Education level | Group | No. (%) | Odds ratio<br>(95% CI) | P value |
|---|---|---|---|---|
| Basic education | General population | 387,476 (18.5%) | 1 [Reference] |  |
|  | Less severe AN | 1050 (18.9%) | **1.60 (1.46-1.74)** | <.001 |
|  | Severe AN | 371 (25.5%) | **2.63 (2.26-3.06)** | <.001 |
| Medium education | General population | 920,322 (43.9%) | [Reference category] |  |
|  | Less severe AN | 1759 (31.6%) | [Reference category] |  |
|  | Severe AN | 448 (30.8%) | [Reference category] |  |
| Short-cycle tertiary and bachelor-level education | General population | 243,498 (11.6%) | 1 [Reference] |  |
|  | Less severe AN | 1009 (18.2%) | **1.21 (1.11-1.32)** | <.001 |
|  | Severe AN | 406 (27.9%) | 0.83 (0.70-0.99) | .042 |
| Master's and doctoral education | General population | 504,239 (24.1%) | 1 [Reference] |  |
|  | Less severe AN | 1684 (30.3%) | 1.05 (0.97-1.13) | .207 |
|  | Severe AN | 211 (14.5%) | 0.87 (0.75-1.01) | .059 |

AN: anorexia nervosa; CI: confidence interval.

### Socio-occupational functioning trajectories

Figure 1 shows associations between illness severity and unemployment, income insufficiency, and living alone. Both severity groups showed worse functioning than the general population across all three domains. Unemployment remained elevated throughout follow-up in both severity groups but followed distinct trajectories. In the less severe group, unemployment odds peaked in midlife (OR =2.66; 95% CI:2.34-3.04 at age 44) and gradually declined towards population levels by age 55 (OR =1.53; 95% CI:1.09-2.15). In contrast, the severe group showed progressively worsening unemployment across adulthood, with ORs increasing from 3.42 (95% CI:3.03-3.86) at age 30 to 4.65 (95% CI:3.54-6.10) at age 45 and rising further later. Income insufficiency risk also diverged over time: in the severe group, odds of low income were stable from age 30 to age 45 (3.84 [95% CI:3.42-4.31] vs. 3.98 [95% CI:3.08-5.14]) and then increased substantially to 7.51 (95% CI:3.85-14.62) at age 55. For cohabitation, both groups consistently showed a higher likelihood of living alone, with a greater magnitude of effect in severe AN (OR = 2.43 (95% CI:2.17-2.71) at age 30; 3.12 (95%CI:1.60-6.05) at age 55). In the joint multinomial model of labour-market status (employment, disability pension, or other unemployment type), with employment as the reference outcome and individuals without AN as the reference group, disability pension risk was significantly elevated in the severe AN group (OR =14.78; 95% CI:12.39-17.64) at age 30, remaining persistently high across adulthood; this pattern was less pronounced in the less severe group (OR =4.78; 95% CI:4.19-5.45, eFigure 1).

**Figure 1.**
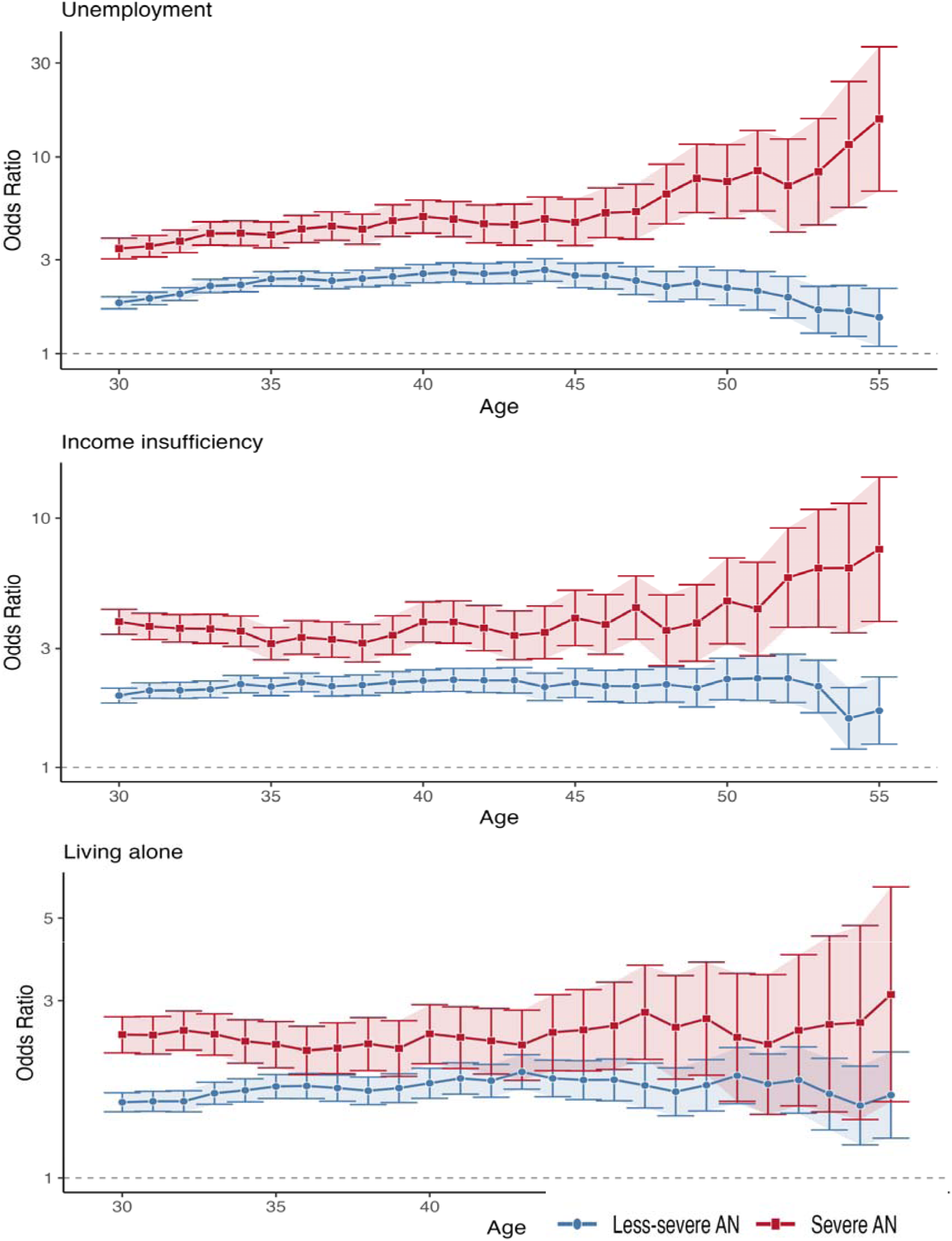
Trajectories of social functioning outcomes in severe and less severe anorexia nervosa relative to the general population. Odds ratios (95% CI) for unemployment, income insufficiency, and living alone among individuals with illness severity groups relative to the general population. Models were adjusted for sex, birth cohort, parental psychiatric history, and parental socioeconomic status. Shaded bands indicate 95% confidence intervals. The dashed horizontal line marks an odds ratio of 1.

### Within-AN severity contrast

Within-AN comparison showed that severity was consistently associated with worse functioning, partly explained by illness status. (Figure 2) Across all three domains, accounting for active illness status marginally attenuated the point estimates toward the null, yet the clear severity gradient persisted independent of current diagnostic status. At age 30, the severe AN group had significantly higher unemployment than the less severe group in the crude model (OR = 1.90; 95% CI:1.65-2.18), which remained robust after adjusting for active AN status (OR = 1.50; 95% CI:1.29-1.74) and active ED status (OR = 1.46; 95% CI:1.26-1.69). A comparable pattern of increased risk held across the follow-up period (ages 30-50), and the severity contrast persisted independent of current diagnostic status (eTable 4). For income insufficiency, the severe-versus-less severe odds at age 30 were attenuated after adjusting for active AN and ED status (OR = 1.98 (95% CI:1.73-2.26); 1.66 (95% CI:1.44-1.93); 1.60 (95% CI:1.38-1.86), respectively). The severity contrast in cohabitation was more modest but followed a similar attenuation pattern (OR = 1.54 (95% CI:1.36-1.75); 1.31 (95% CI:1.14-1.50); 1.29 (95% CI:1.12-1.48), respectively).

**Figure 2.**
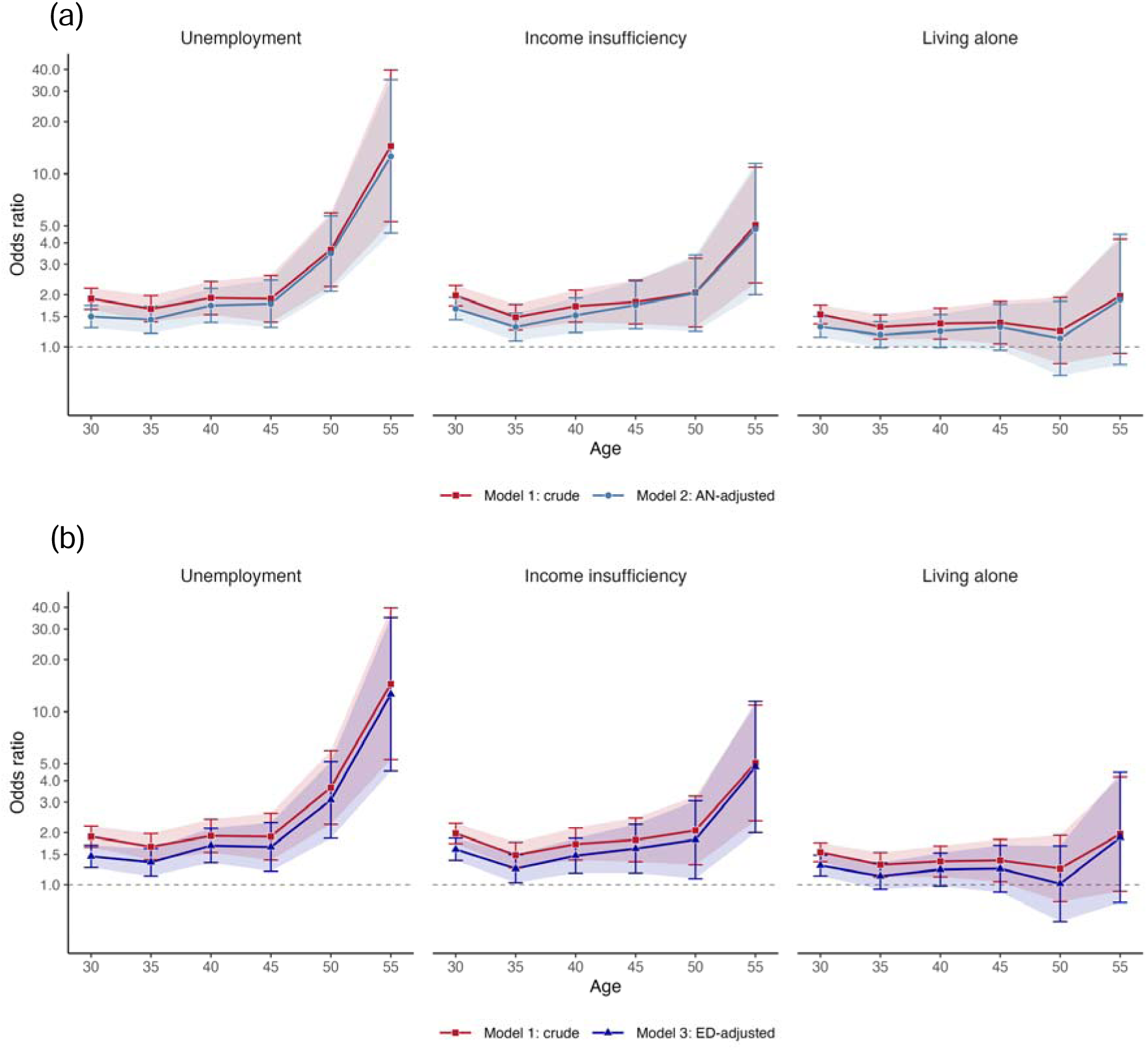
Association between illness severity and social functioning outcomes within individuals with anorexia nervosa. Odds ratios (95% CI) for unemployment, income insufficiency, and living alone among individuals with severe AN relative to those with less severe AN. Model 1 (crude) adjusts for onset age, sex, birth cohort, parental psychiatric history, and parental socioeconomic status. Model 2 (AN-adjusted) additionally adjusts for active AN diagnostic status at the corresponding year. Model 3 (ED-adjusted) additionally adjusts for active ED diagnostic status at the corresponding year. Shaded bands indicate 95% confidence intervals. The dashed horizontal line marks an odds ratio of 1. AN: anorexia nervosa; ED: eating disorders; CI: confidence interval.

### Dose-response analyses

To further examine the dose-response relationship, we evaluated outcomes relative to the general population across AN-RSI severity quintiles (Q1-Q5, from least to most severe). Across all domains, risk estimates followed a similar pattern (Figure 3): ORs were lowest at Q2 and increased monotonically through Q5. For education (not completing upper-secondary), the OR at Q2 was 1.15 (95% CI:0.91-1.45), not significantly different from the general population. For unemployment, income insufficiency, and living alone, ORs remained above 1.0 at every quintile, including the lowest risk point (Q2) (1.35 [95% CI:1.10-1.65], 1.41 [95% CI:1.16-1.72], and 1.37 [95% CI:1.17-1.59], respectively). From Q2 onward, a clear dose-response relationship emerged, with the highest risk for all adverse outcomes concentrated in the most severe quintile (eTable 5).

**Figure 3.**
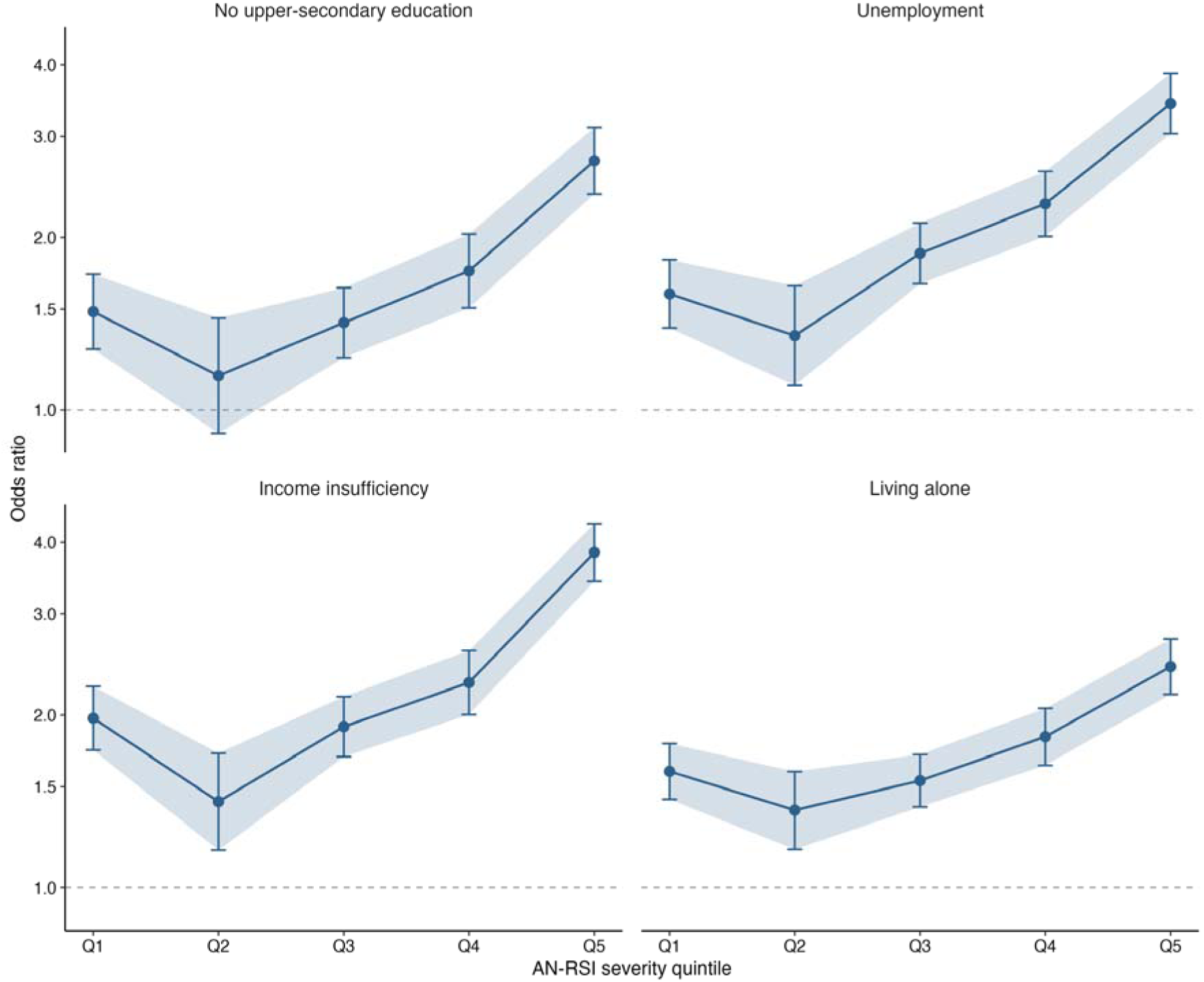
Dose-response relationship between anorexia nervosa illness severity and social functioning outcomes at age 30. Odds ratios (95% CI) for not completing upper secondary education, unemployment, income insufficiency, and living alone at age 30, by AN-RSI severity quintile (Q1, lowest, to Q5, highest), relative to the general population. Error bars indicate 95% confidence intervals. The dashed horizontal line marks an odds ratio of 1. AN-RSI indicates the Anorexia Nervosa Register-based Severity Index; CI, confidence interval.

## Discussion

Utilizing the comprehensive national registers and longitudinal data, we examined a severity-outcome association in AN spanning education, employment, income, and cohabitation. Severe AN carried a substantially greater functioning deficit than less severe AN, and the general population, which persisted—and in some cases widened—over time. Social and occupational impairment observed over time was proportional to illness severity. The association was attenuated, but persisted, after adjustment for concurrent AN or ED hospital contact.

Educational attainment showed a shifting pattern of impairment across ages. By age 25, fewer individuals with AN completed upper-secondary education than the general population. This was more pronounced in the severe AN group and consistent with prior evidence on lower educational attainment in individuals with AN by their late twenties.^[3]^ This deficit coincided with the typical age range for upper-secondary education; severe AN cases had more hospital admissions and longer illness duration, likely disrupting schooling. By age 30 and 35, severe AN cases had a higher likelihood of having completed basic education only and were less likely to have completed higher education. In contrast, less severe AN cases instead showed both increased patterns, at the low and high ends, matching the U-shaped distribution reported in a previous study.^[10]^ This difference may reflect a combination of psychological, cognitive, and familial factors. First, premorbid perfectionism and high achievement orientation may support continued academic engagement among those with less severe AN once they are in remission.^[31]^ In severe AN, the same traits may be present, but their academic advantages may also be attenuated by illness severity and prolonged disruption to continuous education. Second, these academic advantages have been linked to genetic factors associated with AN risk.^[32, 33]^ Individuals with less severe AN may achieve educational levels closer to their familial and genetic potential, whereas greater illness severity may override the advantages. Third, higher familial socioeconomic status may offset the disruption caused by adolescent illness and support return to the educational track.^[8, 34]^ Together, these factors may explain how illness severity shapes long-term educational outcomes.

In line with previous studies^[3, 4]^, individuals with AN had poorer functioning across labour-market outcomes at younger ages, regardless of illness severity. However, this pattern diverged over time: the less severe group became increasingly comparable to the general population, whereas the individuals with severe AN experienced persistent and progressively widening impairment with age. This divergence may explain the inconsistent findings on long-term labour-market outcomes in AN.^[3, 4, 11, 35]^ Prior studies have generally examined AN as a single group and did not account for illness severity. Community-ascertained AN samples,^[11]^ often identified through self-report questionnaires, are more likely to include untreated individuals with mild to moderate illness, which may explain their more favourable outcomes. Clinical cohorts, by contrast, tend to capture individuals with greater severity, longer illness duration, and higher psychiatric comorbidity, resulting in poorer reported outcomes.^[35]^ The widening gap over time points to a process of cumulative disadvantage. Individuals with less severe AN may experience the illness as a temporary disruption; one possibility is that once their illness stabilizes, they may be able to draw upon personal and social support to return to work, eventually catching up to the general population. Conversely, for those with severe AN, prolonged illness duration and repeated treatment or hospitalization often occur during critical periods for entering higher education and building early-career employment, and missing these developmental milestones could potentially result in limited skill development, work experience, and income accumulation, making it increasingly difficult to catch up over time. These divergences suggest that treating AN as a single diagnostic category may mask these severity-dependent differences in long-term functional recovery.^[36]^

When adjusted for hospital contacts of active AN or ED, the association between severe AN and labour-market outcomes was only slightly attenuated at younger ages, with further minimal attenuation at older ages. This suggests that the contribution of concurrent AN/ED illness activity declined steadily across the observation period. The gaps likely reflect a divergence between clinical diagnostic remission and functional recovery.^[37, 38]^ In register-based settings, discharge from active diagnostic status is commonly driven by physical improvement and symptomatic remission.

Using the absence of hospital contact as a proxy for illness remission, our longitudinal data, however, suggest that functional recovery progresses independent of core ED symptoms; diagnostic remission does not appear to reverse the functional vulnerability quickly. This persistence may be compounded by a high burden of psychiatric comorbidity, as previously reported in our AN-RSI validation study.^[17]^ Even after an individual no longer has an ED diagnosis, frequent and long-lasting comorbid psychiatric conditions likely continue to drive functional impairment.^[39]^ Baseline familial disadvantage is an unlikely explanation for this persistence: both severity groups had higher parental socioeconomic status than the general population, which is more pronounced in the severe group. Instead, the persistent impairment is better explained by the timing of AN onset: median AN age at onset is mid-adolescence, a critical period for rapid neurodevelopment and maturation of cortical and subcortical networks.^[40]^ Chronic malnutrition compromises cognitive development, including executive functioning, cognitive flexibility, and decision-making.^[19]^ This developmental stage also coincides with key transitions in education, identity formation, and social development. More severe illness likely reflects greater neurodevelopmental vulnerability, disrupting these transitions and leading to long-term impairment that persists even after remission.

We observed that the severity gradient followed a monotonic pattern, with outcomes worsening consistently at each higher severity level, rather than reflecting only a difference between two broad categories. Functioning outcomes declined stepwise among those with the top 80% scores, consistent with a dose-response relationship. This pattern aligns with the findings from the AN-RSI development study, where increasing AN-RSI scores were associated with a graded increase in mortality risk, supporting the index’s internal construct validity and capacity to capture severity along a continuous gradient rather than a binary distinction.^[17]^

Clinical studies have shown that individual ED symptom severity directly predicts the trajectory of socio-occupational disability and health-related quality of life,^[5]^ and our findings extend this into a population-based setting and demonstrate that the association between clinical severity and functional impairment is not transient but a persistent impact on life-course trajectory. Conversely, the outlier pattern observed at the bottom 20% is most plausibly explained by heterogeneity within the lowest register-severity group.

This group likely comprises both genuinely mild cases and individuals with limited health care contact, potentially reflecting unwillingness to seek treatment—which is not uncommon in AN given its ego-syntonic nature^[41]^—or other unmeasured factors such as later differential diagnosis. This heterogeneity limits interpretation of the bottom 20% as a true mild-illness reference category.

## Strengths and Limitations

Our study benefits from register-based data from Denmark, which provide detailed longitudinal information on health and societal outcomes for all individuals, allowing us to study multiple functional domains over more than 25 years without self-report bias and with minimal loss to follow-up. Another major strength is the use of AN-RSI to measure illness severity, both in terms of binary and gradient effects. We used complementary comparison groups to quantify the absolute burden of AN and isolate the relative effect of illness severity itself. Our nested adjustment strategy allowed us to assess whether the association with illness severity persisted after accounting for current illness status, approximated by register-recorded active AN and ED hospital contacts. This study also has limitations that warrant caution. First, with register-based measurement of illness severity, as discussed above, the heterogeneity within the bottom 20% for AN severity complicates the interpretation of some of the findings in the gradient analysis. Second, due to data availability, we cannot measure clinical presentation and other factors related to AN risk and severity at baseline and at the severity assessment time window, leaving unmeasured or residual confounding factors to consider. Third, this observational study design precludes causal inference. The graded association observed across severity quintiles may indicate that illness severity contributes to observed functional impairment but does not confirm it. Finally, the generalizability of this study may be limited due to differences in clinical practices and welfare systems; validation in other countries is warranted.

## Conclusions

Individuals with lifetime severe AN, regardless of current illness status, showed impaired functioning across multiple domains and increasingly poorer labour-market outcomes across the entire work-life course, indicating a pattern of accumulation rather than static disadvantages. Individuals with less severe illness tend to approximate the general population over time, showing a promising chance of long-term socio-occupational recovery. These findings indicate that register-defined remission does not necessarily correspond to functional recovery. Continued educational and vocational support and monitoring are needed beyond clinical treatment, especially for individuals with a more severe AN course.

## Author contributions

Zeynep Yilmaz served as Principal Investigator and acts as the guarantor of the work. She accepts full responsibility for the integrity of the data, the accuracy of the analyses, and the decision to publish. Study concept and design: All authors. Acquisition, analysis, or interpretation of data: Zi-Ping Zhang, Janne Tidselbak Larsen, Loa Clausen, Esben Agerbo, Cynthia M. Bulik, Liselotte Vogdrup Petersen, and Zeynep Yilmaz. Drafting of the manuscript: Zi-Ping Zhang and Zeynep Yilmaz. Critical revision of the manuscript for important intellectual content: All authors. Statistical analysis: Zi-Ping Zhang.

## Data availability

Data for this study were obtained from Danish national registries via Statistics Denmark and the Danish Health Data Authority. Individual-level data cannot be shared due to Danish data protection legislation and registry access agreements. Qualified researchers may apply for data access through the appropriate Danish authorities, subject to approval of research protocols and compliance with data protection requirements. Statistical analysis code is available from the corresponding author upon request.

## Ethics approval

This study was approved by the Danish Data Protection Agency ( “AU fællesanmeldelse” journal number 2016-051-000001 serial number 2277) and the Danish Health Data Authority. According to Danish law, review by an ethics board and patient consent are not required for purely register-based studies. All data were de-identified and not recognizable at an individual level.

