## Supplementary materials for "Association Between Anorexia Nervosa Severity and Social and Occupational Functioning across Adulthood"

**eMethods**

### **Danish National Registers used in this study**

The Danish Civil Registration System (*Det Centrale Personregister* in Danish, CPR) has recorded basic demographic information on all persons alive and residing in Denmark since 1968.^[21]^ It captures vital status, dates of birth, death, and emigration. Every Danish resident is assigned a unique personal identification number, which enables accurate linkage across all Danish national registers.

The Danish National Patient Register and the Danish Psychiatric Central Research Register together record all psychiatric inpatient admissions since 1969 and all outpatient contacts since 1995, covering the full Danish population.^[22, 23]^ Diagnoses are coded according to ICD-8 (until 1994) and ICD-10 (from 1995 onward). Anorexia nervosa (AN) and other eating disorders diagnoses, as well as psychiatric comorbidities, were ascertained from these two registers.

Educational attainment was obtained from the Population's Education Register (PER), which is the most widely used of several Danish education registers.^[24]^ Since 2000, the PER has recorded the highest completed education level annually for individuals aged 15 to 69, covering over 96.4% of the Danish population.^[24]^

Labor market and income data were obtained from the Employment Classification Module (AKM) and the Integrated Database for Labor Market Research (IDA).^[25]^ Since 1980, AKM has provided annually updated information on employment status and occupational classification. IDA integrates longitudinal individual-level data on labor market participation and income.

Cohabitation status data were obtained from the Population Statistics Register (*Befolkningen* in Danish, BEF).^[26]^ Since 1971, this register has provided information on births, deaths, relocations, emigration/immigration, marriage-related events, etc., from the CPR.

All registers cover the full Danish population.

### **Illness Severity of Anorexia Nervosa**

Illness severity was quantified within the AN cohort using the AN Register-based Severity Index (AN-RSI), a register-based measure developed by Larsen et al.^[17]^ The AN-RSI is calculated as the sum of weighted points assigned to six severity-related variables: early onset (defined as age below 10 years at first AN treatment; 1 point), late onset (defined as age above 25 years; 1 point), number of inpatient admissions (2 points per admission), number of outpatient re-contacts beyond the first treatment course (1 point per contact), cumulative treatment length (1 point per full year), and illness duration defined as time from first diagnosis to last treatment within 5 years post-diagnosis (1 point per full year). Full details of the weighting procedure are described in Larsen et al.^[17]^ All variables were derived from the Danish National Patient Register and the Danish Psychiatric Central Research Register. Since the present study includes only individuals with a first AN diagnosis between ages 10 and 25, both the early onset and late onset components were not applicable and were omitted from the AN-RSI calculation. The AN-RSI was evaluated 5 years after the first AN diagnosis, consistent with the original paper, reflecting the intermediate illness duration criterion used to identify severe AN.^[15, 17, 41]^

Individuals were classified into severity quintiles based on the AN-RSI distribution in the full AN cohort. For binary classification, those scoring in the top 20% were designated as severe AN and the remainder as less-severe AN, which corresponds to the proportion of individuals with AN who do not improve with treatment and is consistent with the threshold used in Larsen et al.^[2, 17, 42, 43]^

### **Outcomes**

Outcome definitions and coding mainly followed the SEPLINE guidelines for socioeconomic position measurement using Danish register data.^[29]^

Educational outcomes were measured at ages 25 and 30. Education level at a specific age point was derived from Statistics Denmark's DISCED-15 classification, harmonized with International Standard Classification of Education (ISCED) 2011.^[44]^ In Denmark, upper secondary education corresponds to ISCED level 3 and includes both general and vocational programs, typically commencing at the end of full-time compulsory schooling. Completion of upper secondary education was assessed as a binary variable (yes/no) at age 25. The highest completed education level was assessed at age 30 and 35. This age was selected for two reasons: first, the majority of the Danish population has attained their highest education level by age 30 ^[29]^, and individuals require a further 5 years to have their AN-RSI calculated, making age 35 the feasible measurement point for the full cohort and ensuring most individuals finish their education. Age 30 was included as a sensitivity analysis to capture earlier completers while remaining close to the primary assessment window, given the oldest age of onset was set to 25 in our cohort. The highest education level was categorized into four ordered levels: (1) Level 1: basic education (ISCED 0-2); (2) Level 2: medium education (including upper secondary education and post–secondary non-tertiary education, ISCED 3-4); (3) Level 3: short-cycle tertiary and bachelor-level education (ISCED 5-6); and (4) Level 4: master's and doctoral education (ISCED 7-8).

Employment status was captured from AKM and IDA and coded as a binary variable: employment (including working and enrolled in education) versus unemployment (including temporary unemployment, retirement, and individuals outside the workforce, such as those receiving social security, sickness benefits, or disability pension). As a secondary outcome for employment status, this outcome was further coded into three categories: employment, receiving disability pension, and other unemployment.

Yearly income was derived from annual wage and/or self-employment earnings in a calendar year from AKM and IDA. Income deficiency was defined as placement in the lowest income quintile, relative to the sex- and five-year age group-matched general population in Denmark.

Cohabitation status was derived from the CPR and coded as a binary variable: cohabiting (including married, registered partnership, or cohabiting with a partner) versus not cohabiting (single).

For all binary outcomes, higher odds ratios indicate less favorable social functioning. This corresponds to non-completion of upper-secondary education at age 25, unemployment relative to employment, income deficiency relative to higher income, and living alone relative to cohabiting. For the secondary categorical employment outcome, both non-reference categories in this domain represent less favorable functioning relative to employment; an OR above 1 for either disability pension or other unemployment indicates a less favorable outcome.

For highest educational attainment at age 30, coded as an ordinal four-level outcome with medium education (Level 2) as the reference category, direction depends on the comparison level rather than following a single rule. An OR above 1 for Level 1 (basic education) indicates a less favorable outcome, while an OR above 1 for Level 3 (short-cycle tertiary or bachelor-level) or Level 4 (master's or doctoral) indicates a more favorable outcome. This reflects the ordinal structure of educational attainment rather than a binary unfavorable/favorable classification.

**Covariates**

All models were adjusted for a set of covariates, including sex, birth year, and age of onset (for within-AN comparison only), parental psychiatric history, and parental unfavorable socioeconomic position indicators. Linkage of parents was derived from the CPR. The history of parental psychiatric disorder was obtained from health registers and coded as any versus no parental psychiatric diagnosis. Unfavorable parental socioeconomic position indicators were assessed when participants were aged 25 years, using register information on employment status (unemployed/employed), income (income deficiency or not), and the highest completed education (no completion of upper-secondary education) for both parents. These three variables were entered as separate binary covariates, rather than combined into a single composite score.

26.	*Danish Population Statistics*.

44.	OECD, E., UNESCO Institute for Statistics, *ISCED 2011 Operational Manual: Guidelines for Classifying National Education Programmes and Related Qualifications.* OECD Publishing, 2015.

**eFigure 1. Trajectories of employment status in severe and less-severe anorexia nervosa relative to the general population (employment as reference category)**

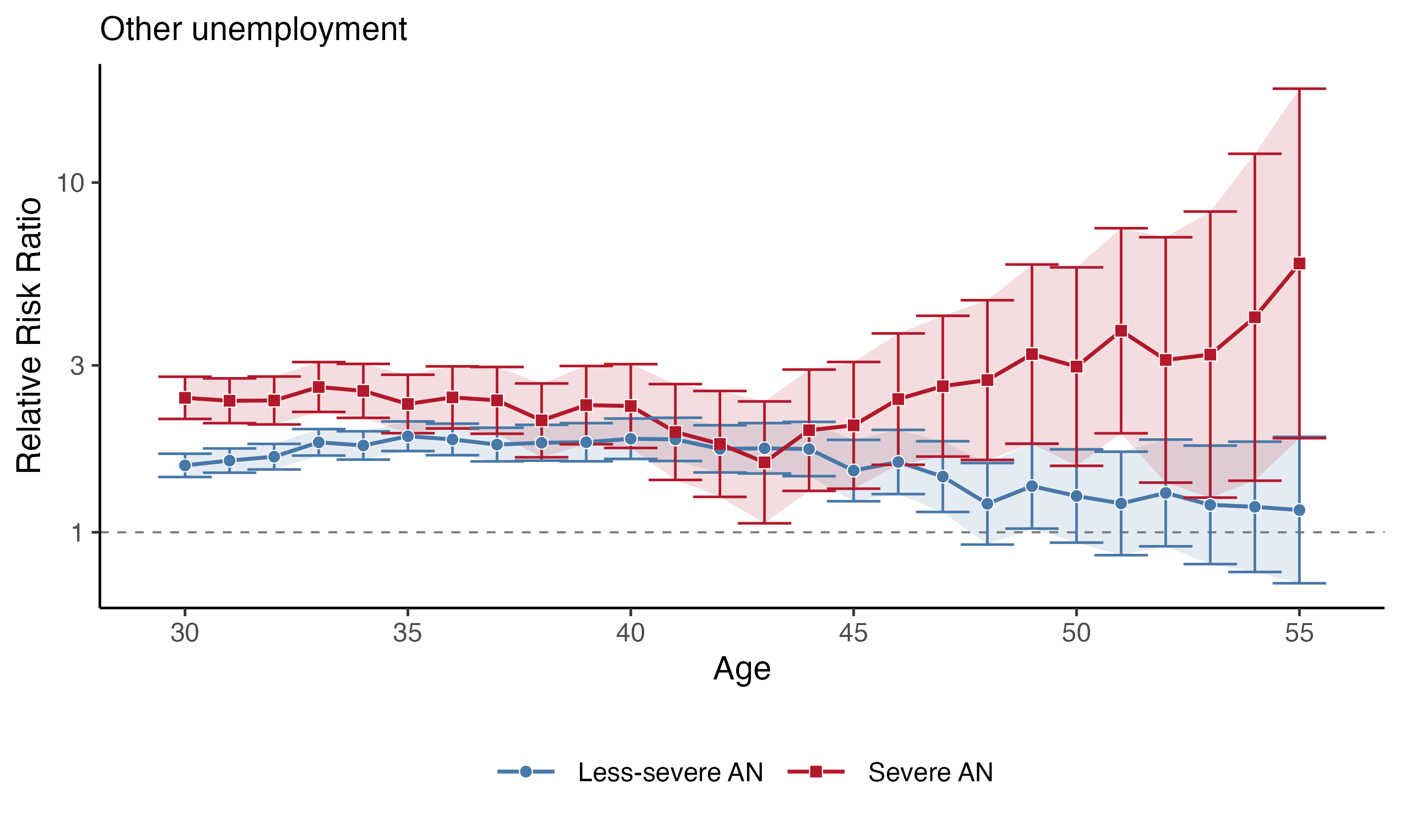

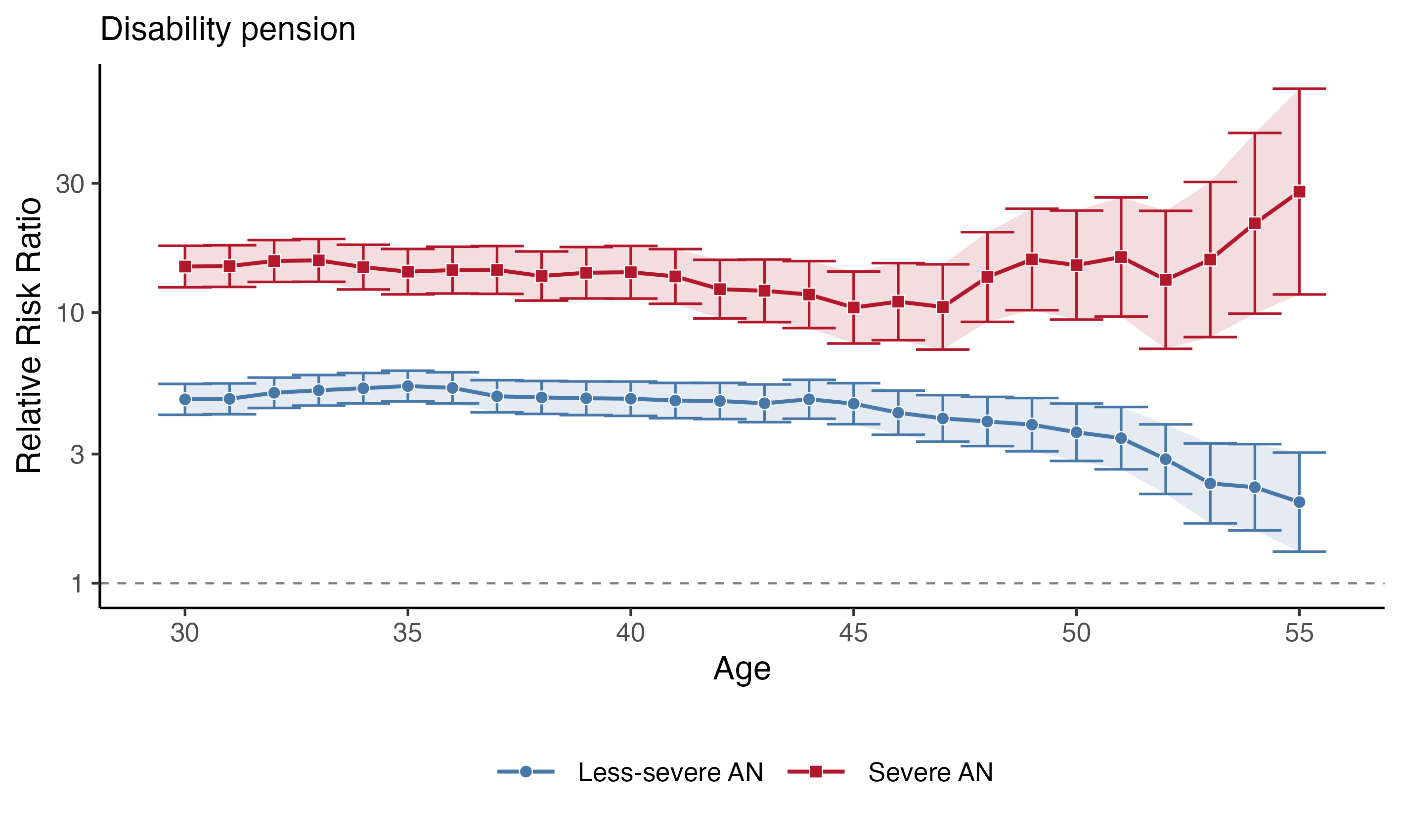

***Legend****:* Odds ratios (95% CI) for receiving disability pension and other unemployment (employment as reference category) among individuals with illness severity groups against the general population. Models were adjusted for sex, birth cohort, parental psychiatric history, and parental socioeconomic status. Shaded bands indicate 95% confidence intervals. The dashed horizontal line marks an odds ratio of 1.

**eTable 1.** Number of Individuals With Anorexia Nervosa and in the General Population Contributing at Each Age of Follow-up

|  | **2024** | | | **2023** | | |
| --- | --- | --- | --- | --- | --- | --- |
| **Age** | **Less-severe AN** | **Severe AN** | **General population** | **Less-severe AN** | **Severe AN** | **General population** |
| 25 | 5219 | 1467 | 2432451 |  |  |  |
| 30 | 5469 | 1442 | 2032009 | 5143 | 1345 | 1994261 |
| 31 | 5075 | 1335 | 1961997 | 4784 | 1252 | 1927865 |
| 32 | 4699 | 1234 | 1893545 | 4404 | 1147 | 1859748 |
| 33 | 4302 | 1131 | 1825881 | 4054 | 1054 | 1794099 |
| 34 | 3963 | 1039 | 1761575 | 3720 | 942 | 1729252 |
| 35 | 3650 | 930 | 1697989 | 3409 | 871 | 1666831 |
| 36 | 3345 | 859 | 1636826 | 3137 | 784 | 1606856 |
| 37 | 3082 | 771 | 1578119 | 2887 | 716 | 1549513 |
| 38 | 2831 | 706 | 1522188 | 2626 | 652 | 1493085 |
| 39 | 2583 | 637 | 1466968 | 2398 | 581 | 1438349 |
| 40 | 2360 | 569 | 1412829 | 2187 | 531 | 1385637 |
| 41 | 2145 | 520 | 1360879 | 1983 | 476 | 1334038 |
| 42 | 1950 | 463 | 1310126 | 1792 | 413 | 1280913 |
| 43 | 1764 | 402 | 1257855 | 1609 | 355 | 1227646 |
| 44 | 1588 | 349 | 1205462 | 1438 | 313 | 1170634 |
| 45 | 1412 | 305 | 1149281 | 1255 | 272 | 1111906 |
| 46 | 1231 | 264 | 1091482 | 1100 | 231 | 1050693 |
| 47 | 1077 | 223 | 1031359 | 972 | 198 | 989582 |
| 48 | 946 | 191 | 971472 | 855 | 166 | 925483 |
| 49 | 837 | 162 | 908393 | 738 | 135 | 855674 |
| 50 | 721 | 132 | 839739 | 649 | 117 | 786972 |
| 51 | 632 | 113 | 771940 | 564 | 95 | 718276 |
| 52 | 547 | 91 | 704394 | 503 | 74 | 646388 |
| 53 | 484 | 72 | 633658 | 434 | 61 | 575083 |
| 54 | 421 | 59 | 563629 | 380 | 52 | 507866 |
| 55 | 365 | 51 | 497446 | 324 | 46 | 441132 |

Individual counts are shown separately for the 2023 and 2024 Danish register extracts. Age 25 reflects the education outcome only; ages 26-29 were not examined.

**eTable 2.** Number of Outcome Events for Employment, Income, and Cohabitation Status by Age and Anorexia Nervosa Severity, Relative to the General Population

|  | **Employment** | | | | | | **Income** | | | | | | **Cohabitation** | | | | | |
| --- | --- | --- | --- | --- | --- | --- | --- | --- | --- | --- | --- | --- | --- | --- | --- | --- | --- | --- |
|  | **General population** | | **Less severe AN** | | **Severe AN** | | **General population** | | **Less severe AN** | | **Severe AN** | | **General population** | | **Less severe AN** | | **Severe AN** | |
| **age** | **unemployed** | **employed** | **unemployed** | **employed** | **unemployed** | **employed** | **Low level** | **Medium and high level** | **Low level** | **Medium and high level** | **Low level** | **Medium and high level** | **Living alone** | **Not living alone** | **Living alone** | **Not living alone** | **Living alone** | **Not living alone** |
| 30 | 309424 (15.5%) | 1684837 (84.5%) | 1368 (26.6%) | 3775 (73.4%) | 519 (38.6%) | 826 (61.4%) | 317163 (15.4%) | 1745527 (84.6%) | 1391 (25.1%) | 4142 (74.9%) | 549 (37.9%) | 898 (62.1%) | 686293 (33.8%) | 1345716 (66.2%) | 2235 (40.9%) | 3234 (59.1%) | 737 (51.1%) | 705 (48.9%) |
| 31 | 293874 (15.2%) | 1633991 (84.8%) | 1302 (27.2%) | 3482 (72.8%) | 488 (39.0%) | 764 (61.0%) | 301454 (15.1%) | 1692538 (84.9%) | 1301 (25.3%) | 3849 (74.7%) | 491 (36.5%) | 853 (63.5%) | 620072 (31.6%) | 1341925 (68.4%) | 1976 (38.9%) | 3099 (61.1%) | 655 (49.1%) | 680 (50.9%) |
| 32 | 275444 (14.8%) | 1584304 (85.2%) | 1216 (27.6%) | 3188 (72.4%) | 453 (39.5%) | 694 (60.5%) | 288151 (15.0%) | 1638164 (85.0%) | 1187 (24.9%) | 3585 (75.1%) | 448 (35.9%) | 801 (64.1%) | 566411 (29.9%) | 1327134 (70.1%) | 1757 (37.4%) | 2942 (62.6%) | 593 (48.1%) | 641 (51.9%) |
| 33 | 258747 (14.4%) | 1535352 (85.6%) | 1170 (28.9%) | 2884 (71.1%) | 424 (40.2%) | 630 (59.8%) | 274840 (14.8%) | 1582661 (85.2%) | 1083 (24.6%) | 3311 (75.4%) | 395 (34.5%) | 750 (65.5%) | 522899 (28.6%) | 1302982 (71.4%) | 1593 (37.0%) | 2709 (63.0%) | 523 (46.2%) | 608 (53.8%) |
| 34 | 243011 (14.1%) | 1486241 (85.9%) | 1052 (28.3%) | 2668 (71.7%) | 376 (39.9%) | 566 (60.1%) | 262815 (14.7%) | 1529313 (85.3%) | 1022 (25.3%) | 3011 (74.7%) | 348 (33.2%) | 701 (66.8%) | 488006 (27.7%) | 1273569 (72.3%) | 1459 (36.8%) | 2504 (63.2%) | 459 (44.2%) | 580 (55.8%) |
| 35 | 230415 (13.8%) | 1436416 (86.2%) | 988 (29.0%) | 2421 (71.0%) | 335 (38.5%) | 536 (61.5%) | 266051 (15.4%) | 1461490 (84.6%) | 993 (26.8%) | 2718 (73.2%) | 321 (34.1%) | 620 (65.9%) | 459025 (27.0%) | 1238964 (73.0%) | 1331 (36.5%) | 2319 (63.5%) | 404 (43.4%) | 526 (56.6%) |
| 36 | 219814 (13.7%) | 1387042 (86.3%) | 904 (28.8%) | 2233 (71.2%) | 315 (40.2%) | 469 (59.8%) | 256253 (15.4%) | 1408858 (84.6%) | 920 (27.1%) | 2480 (72.9%) | 301 (34.7%) | 567 (65.3%) | 435928 (26.6%) | 1200898 (73.4%) | 1220 (36.5%) | 2125 (63.5%) | 363 (42.3%) | 496 (57.7%) |
| 37 | 210462 (13.6%) | 1339051 (86.4%) | 820 (28.4%) | 2067 (71.6%) | 290 (40.5%) | 426 (59.5%) | 247306 (15.4%) | 1357873 (84.6%) | 818 (26.1%) | 2316 (73.9%) | 267 (34.2%) | 514 (65.8%) | 417892 (26.5%) | 1160227 (73.5%) | 1125 (36.5%) | 1957 (63.5%) | 328 (42.5%) | 443 (57.5%) |
| 38 | 201801 (13.5%) | 1291284 (86.5%) | 753 (28.7%) | 1873 (71.3%) | 259 (39.7%) | 393 (60.3%) | 238680 (15.4%) | 1309228 (84.6%) | 761 (26.4%) | 2120 (73.6%) | 240 (33.5%) | 476 (66.5%) | 403206 (26.5%) | 1118982 (73.5%) | 1039 (36.7%) | 1792 (63.3%) | 303 (42.9%) | 403 (57.1%) |
| 39 | 194276 (13.5%) | 1244073 (86.5%) | 681 (28.4%) | 1717 (71.6%) | 242 (41.7%) | 339 (58.3%) | 230872 (15.5%) | 1260908 (84.5%) | 703 (26.9%) | 1915 (73.1%) | 229 (35.2%) | 421 (64.8%) | 390881 (26.6%) | 1076087 (73.4%) | 966 (37.4%) | 1617 (62.6%) | 277 (43.5%) | 360 (56.5%) |
| 40 | 187615 (13.5%) | 1198022 (86.5%) | 634 (29.0%) | 1553 (71.0%) | 228 (42.9%) | 303 (57.1%) | 224788 (15.6%) | 1212272 (84.4%) | 684 (28.5%) | 1716 (71.5%) | 223 (38.6%) | 354 (61.4%) | 378484 (26.8%) | 1034345 (73.2%) | 909 (38.5%) | 1451 (61.5%) | 261 (45.9%) | 308 (54.1%) |
| 41 | 180025 (13.5%) | 1154013 (86.5%) | 576 (29.0%) | 1407 (71.0%) | 203 (42.6%) | 273 (57.4%) | 217460 (15.7%) | 1166867 (84.3%) | 605 (27.7%) | 1576 (72.3%) | 212 (40.0%) | 318 (60.0%) | 369228 (27.1%) | 991651 (72.9%) | 851 (39.7%) | 1294 (60.3%) | 240 (46.2%) | 280 (53.8%) |
| 42 | 173231 (13.5%) | 1107682 (86.5%) | 522 (29.1%) | 1270 (70.9%) | 176 (42.6%) | 237 (57.4%) | 210880 (15.8%) | 1121803 (84.2%) | 557 (28.1%) | 1422 (71.9%) | 185 (39.0%) | 289 (61.0%) | 360188 (27.5%) | 949938 (72.5%) | 770 (39.5%) | 1180 (60.5%) | 217 (46.9%) | 246 (53.1%) |
| 43 | 167151 (13.6%) | 1060495 (86.4%) | 473 (29.4%) | 1136 (70.6%) | 152 (42.8%) | 203 (57.2%) | 203802 (15.9%) | 1075760 (84.1%) | 503 (28.2%) | 1283 (71.8%) | 152 (37.3%) | 256 (62.7%) | 351348 (27.9%) | 906507 (72.1%) | 728 (41.3%) | 1036 (58.7%) | 190 (47.3%) | 212 (52.7%) |
| 44 | 161824 (13.8%) | 1008810 (86.2%) | 427 (29.7%) | 1011 (70.3%) | 138 (44.1%) | 175 (55.9%) | 197455 (16.1%) | 1028910 (83.9%) | 432 (26.8%) | 1177 (73.2%) | 136 (38.4%) | 218 (61.6%) | 341853 (28.4%) | 863609 (71.6%) | 657 (41.4%) | 931 (58.6%) | 175 (50.1%) | 174 (49.9%) |
| 45 | 157258 (14.1%) | 954648 (85.9%) | 371 (29.6%) | 884 (70.4%) | 116 (42.6%) | 156 (57.4%) | 182895 (15.6%) | 986236 (84.4%) | 382 (26.7%) | 1049 (73.3%) | 126 (40.4%) | 186 (59.6%) | 331159 (28.8%) | 818122 (71.2%) | 592 (41.9%) | 820 (58.1%) | 155 (50.8%) | 150 (49.2%) |
| 46 | 152629 (14.5%) | 898064 (85.5%) | 322 (29.3%) | 778 (70.7%) | 104 (45.0%) | 127 (55.0%) | 176233 (15.9%) | 934050 (84.1%) | 338 (27.0%) | 916 (73.0%) | 107 (39.6%) | 163 (60.4%) | 318999 (29.2%) | 772483 (70.8%) | 520 (42.2%) | 711 (57.8%) | 139 (52.7%) | 125 (47.3%) |
| 47 | 146515 (14.8%) | 843067 (85.2%) | 277 (28.5%) | 695 (71.5%) | 90 (45.5%) | 108 (54.5%) | 169216 (16.1%) | 879748 (83.9%) | 294 (26.8%) | 804 (73.2%) | 98 (42.6%) | 132 (57.4%) | 305479 (29.6%) | 725880 (70.4%) | 448 (41.6%) | 629 (58.4%) | 120 (53.8%) | 103 (46.2%) |
| 48 | 139321 (15.1%) | 786162 (84.9%) | 240 (28.1%) | 615 (71.9%) | 82 (49.4%) | 84 (50.6%) | 161471 (16.3%) | 826324 (83.7%) | 259 (26.8%) | 707 (73.2%) | 75 (38.3%) | 121 (61.7%) | 290427 (29.9%) | 681045 (70.1%) | 388 (41.0%) | 558 (59.0%) | 100 (52.4%) | 91 (47.6%) |
| 49 | 131316 (15.3%) | 724358 (84.7%) | 217 (29.4%) | 521 (70.6%) | 71 (52.6%) | 64 (47.4%) | 153745 (16.6%) | 770030 (83.4%) | 231 (27.0%) | 623 (73.0%) | 66 (40.0%) | 99 (60.0%) | 273736 (30.1%) | 634657 (69.9%) | 358 (42.8%) | 479 (57.2%) | 86 (53.1%) | 76 (46.9%) |
| 50 | 123625 (15.7%) | 663347 (84.3%) | 187 (28.8%) | 462 (71.2%) | 61 (52.1%) | 56 (47.9%) | 133185 (15.6%) | 720894 (84.4%) | 199 (27.0%) | 538 (73.0%) | 56 (41.8%) | 78 (58.2%) | 254535 (30.3%) | 585204 (69.7%) | 318 (44.1%) | 403 (55.9%) | 67 (50.8%) | 65 (49.2%) |
| 51 | 115186 (16.0%) | 603090 (84.0%) | 158 (28.0%) | 406 (72.0%) | 57 (60.0%) | 38 (40.0%) | 124985 (15.9%) | 660366 (84.1%) | 174 (26.9%) | 472 (73.1%) | 48 (41.4%) | 68 (58.6%) | 235489 (30.5%) | 536451 (69.5%) | 272 (43.0%) | 360 (57.0%) | 57 (50.4%) | 56 (49.6%) |
| 52 | 106565 (16.5%) | 539823 (83.5%) | 141 (28.0%) | 362 (72.0%) | 43 (58.1%) | 31 (41.9%) | 116226 (16.2%) | 600440 (83.8%) | 151 (26.8%) | 412 (73.2%) | 46 (48.9%) | 48 (51.1%) | 215889 (30.6%) | 488505 (69.4%) | 239 (43.7%) | 308 (56.3%) | 48 (52.7%) | 43 (47.3%) |
| 53 | 97658 (17.0%) | 477425 (83.0%) | 116 (26.7%) | 318 (73.3%) | 37 (60.7%) | 24 (39.3%) | 107326 (16.6%) | 537377 (83.4%) | 133 (26.7%) | 366 (73.3%) | 37 (51.4%) | 35 (48.6%) | 195134 (30.8%) | 438524 (69.2%) | 205 (42.4%) | 279 (57.6%) | 38 (52.8%) | 34 (47.2%) |
| 54 | 88620 (17.4%) | 419246 (82.6%) | 103 (27.1%) | 277 (72.9%) | 34 (65.4%) | 18 (34.6%) | 98080 (17.1%) | 475522 (82.9%) | 101 (23.4%) | 330 (76.6%) | 33 (54.1%) | 28 (45.9%) | 174280 (30.9%) | 389349 (69.1%) | 177 (42.0%) | 244 (58.0%) | 30 (50.8%) | 29 (49.2%) |
| 55 | 79530 (18.0%) | 361602 (82.0%) | 88 (27.2%) | 236 (72.8%) | 32 (69.6%) | 14 (30.4%) | 82866 (16.4%) | 423603 (83.6%) | 86 (22.9%) | 290 (77.1%) | 29 (55.8%) | 23 (44.2%) | 154473 (31.1%) | 342973 (68.9%) | 158 (43.3%) | 207 (56.7%) | 27 (52.9%) | 24 (47.1%) |

**eTable 3.** Relative Risk of Highest Educational Level by Anorexia Nervosa Severity at Age 30

| **Education level** | **Group** | **No. (%)** | **Odds ratio** | **P value** |
| --- | --- | --- | --- | --- |
|  |  |  | **(95% CI)** |  |
| **Age 35** | | | | |
| Basic education | General population | 297,651 (16.9%) | 1 [Reference] |  |
|  | Less-severe AN | 663 (17.6%) | 1.77 (1.58-1.98) | <.001 |
|  | Severe AN | 220 (23.0%) | 3.02 (2.47-3.69) | <.001 |
| Medium education | General population | 750,467 (42.5%) | [Reference category] |  |
|  | Less-severe AN | 1120 (29.7%) | [Reference category] |  |
|  | Severe AN | 256 (26.8%) | [Reference category] |  |
| Short-cycle tertiary and bachelor-level education | General population | 241,948 (13.7%) | 1 [Reference] |  |
|  | Less-severe AN | 814 (21.6%) | 1.40 (1.26-1.55) | <.001 |
|  | Severe AN | 173 (18.1%) | 1.12 (0.90-1.38) | .305 |
| Master's and doctoral education | General population | 441,182 (25.0%) | 1 [Reference] |  |
|  | Less-severe AN | 1128 (30.0%) | 1.10 (1.00-1.21) | .043 |
|  | Severe AN | 294 (30.8%) | 1.09 (0.91-1.32) | .337 |

AN, anorexia nervosa; CI, confidence interval.

**eTable 4.** Association Between Illness Severity and Employment, Income, and Cohabitation Outcomes Within Individuals With Anorexia Nervosa, by Age

|  | **Work** | | | | | | **Income** | | | | | | **Cohabitation** | | | | | |
| --- | --- | --- | --- | --- | --- | --- | --- | --- | --- | --- | --- | --- | --- | --- | --- | --- | --- | --- |
| **Age** | **Model 1** | | **Model 2** | | **Model 3** | | **Model 1** | | **Model 2** | | **Model 3** | | **Model 1** | | **Model 2** | | **Model 3** | |
|  | **OR (95% CI)** | **P value** | **OR (95% CI)** | **P value** | **OR (95% CI)** | **P value** | **OR (95% CI)** | **P value** | **OR (95% CI)** | **P value** | **OR (95% CI)** | **P value** | **OR (95% CI)** | **P value** | **OR (95% CI)** | **P value** | **OR (95% CI)** | **P value** |
| 30 | 1.90 (1.65-2.18) | <.001 | 1.50 (1.29-1.74) | <.001 | 1.46 (1.26-1.69) | <.001 | 1.98 (1.73-2.26) | <.001 | 1.66 (1.44-1.93) | <.001 | 1.60 (1.38-1.86) | <.001 | 1.54 (1.36-1.75) | <.001 | 1.31 (1.14-1.50) | <.001 | 1.29 (1.12-1.48) | <.001 |
| 31 | 1.85 (1.60-2.14) | <.001 | 1.53 (1.31-1.79) | <.001 | 1.49 (1.28-1.74) | <.001 | 1.81 (1.57-2.08) | <.001 | 1.50 (1.29-1.75) | <.001 | 1.43 (1.23-1.67) | <.001 | 1.54 (1.34-1.75) | <.001 | 1.29 (1.12-1.49) | <.001 | 1.27 (1.10-1.46) | .001 |
| 32 | 1.85 (1.59-2.16) | <.001 | 1.56 (1.33-1.83) | <.001 | 1.51 (1.28-1.77) | <.001 | 1.78 (1.53-2.06) | <.001 | 1.55 (1.32-1.82) | <.001 | 1.50 (1.28-1.76) | <.001 | 1.58 (1.37-1.81) | <.001 | 1.37 (1.18-1.59) | <.001 | 1.31 (1.13-1.53) | <.001 |
| 33 | 1.84 (1.58-2.16) | <.001 | 1.58 (1.34-1.86) | <.001 | 1.49 (1.26-1.75) | <.001 | 1.75 (1.50-2.05) | <.001 | 1.51 (1.28-1.79) | <.001 | 1.45 (1.22-1.72) | <.001 | 1.46 (1.27-1.69) | <.001 | 1.24 (1.06-1.46) | .007 | 1.20 (1.02-1.40) | .027 |
| 34 | 1.83 (1.55-2.16) | <.001 | 1.56 (1.31-1.86) | <.001 | 1.48 (1.24-1.76) | <.001 | 1.63 (1.39-1.92) | <.001 | 1.39 (1.17-1.67) | <.001 | 1.32 (1.10-1.58) | .003 | 1.38 (1.18-1.60) | <.001 | 1.19 (1.00-1.40) | .044 | 1.14 (0.97-1.35) | .118 |
| 35 | 1.66 (1.40-1.98) | <.001 | 1.44 (1.20-1.72) | <.001 | 1.35 (1.12-1.62) | .001 | 1.48 (1.25-1.76) | <.001 | 1.30 (1.08-1.57) | .005 | 1.24 (1.03-1.49) | .025 | 1.30 (1.11-1.53) | .001 | 1.18 (0.99-1.40) | .067 | 1.12 (0.94-1.34) | .198 |
| 36 | 1.77 (1.48-2.13) | <.001 | 1.50 (1.24-1.82) | <.001 | 1.41 (1.17-1.72) | <.001 | 1.52 (1.27-1.82) | <.001 | 1.34 (1.11-1.63) | .003 | 1.28 (1.05-1.55) | .014 | 1.24 (1.04-1.46) | .014 | 1.14 (0.94-1.36) | .176 | 1.10 (0.91-1.32) | .332 |
| 37 | 1.87 (1.54-2.26) | <.001 | 1.59 (1.31-1.94) | <.001 | 1.52 (1.24-1.86) | <.001 | 1.55 (1.28-1.87) | <.001 | 1.35 (1.10-1.66) | .004 | 1.31 (1.06-1.61) | .011 | 1.27 (1.06-1.52) | .008 | 1.12 (0.92-1.36) | .256 | 1.08 (0.89-1.31) | .428 |
| 38 | 1.75 (1.43-2.14) | <.001 | 1.52 (1.23-1.87) | <.001 | 1.43 (1.16-1.77) | <.001 | 1.47 (1.20-1.79) | <.001 | 1.27 (1.02-1.57) | .031 | 1.21 (0.97-1.50) | .091 | 1.32 (1.09-1.59) | .004 | 1.17 (0.95-1.43) | .132 | 1.13 (0.93-1.39) | .225 |
| 39 | 1.90 (1.54-2.35) | <.001 | 1.68 (1.35-2.09) | <.001 | 1.59 (1.27-1.99) | <.001 | 1.54 (1.26-1.89) | <.001 | 1.43 (1.15-1.79) | .002 | 1.35 (1.08-1.69) | .009 | 1.27 (1.05-1.55) | .015 | 1.20 (0.97-1.48) | .098 | 1.17 (0.95-1.45) | .144 |
| 40 | 1.92 (1.54-2.39) | <.001 | 1.74 (1.38-2.18) | <.001 | 1.69 (1.34-2.12) | <.001 | 1.72 (1.39-2.13) | <.001 | 1.52 (1.21-1.92) | <.001 | 1.47 (1.17-1.86) | .001 | 1.36 (1.11-1.68) | .003 | 1.24 (0.99-1.54) | .061 | 1.22 (0.98-1.53) | .074 |
| 41 | 1.85 (1.46-2.33) | <.001 | 1.65 (1.30-2.10) | <.001 | 1.60 (1.25-2.04) | <.001 | 1.70 (1.36-2.12) | <.001 | 1.44 (1.13-1.83) | .003 | 1.40 (1.09-1.79) | .007 | 1.30 (1.05-1.62) | .016 | 1.18 (0.94-1.50) | .158 | 1.15 (0.91-1.45) | .254 |
| 42 | 1.77 (1.37-2.27) | <.001 | 1.62 (1.25-2.09) | <.001 | 1.56 (1.20-2.02) | <.001 | 1.60 (1.26-2.03) | <.001 | 1.39 (1.08-1.80) | .012 | 1.34 (1.03-1.74) | .028 | 1.28 (1.02-1.61) | .032 | 1.08 (0.84-1.39) | .533 | 1.05 (0.82-1.36) | .698 |
| 43 | 1.75 (1.33-2.29) | <.001 | 1.60 (1.21-2.11) | .001 | 1.52 (1.15-2.02) | .004 | 1.50 (1.16-1.94) | .002 | 1.44 (1.09-1.91) | .011 | 1.38 (1.04-1.83) | .027 | 1.20 (0.94-1.53) | .153 | 1.18 (0.90-1.55) | .233 | 1.13 (0.86-1.48) | .398 |
| 44 | 1.82 (1.36-2.42) | <.001 | 1.67 (1.25-2.25) | <.001 | 1.61 (1.20-2.17) | .002 | 1.62 (1.23-2.13) | <.001 | 1.49 (1.10-2.01) | .009 | 1.43 (1.06-1.93) | .021 | 1.36 (1.04-1.77) | .023 | 1.28 (0.96-1.71) | .091 | 1.25 (0.93-1.67) | .133 |
| 45 | 1.90 (1.39-2.59) | <.001 | 1.78 (1.29-2.44) | <.001 | 1.65 (1.20-2.28) | .002 | 1.82 (1.36-2.44) | <.001 | 1.75 (1.27-2.40) | <.001 | 1.62 (1.17-2.24) | .004 | 1.38 (1.04-1.83) | .024 | 1.30 (0.95-1.77) | .096 | 1.24 (0.91-1.69) | .179 |
| 46 | 2.14 (1.53-2.99) | <.001 | 2.03 (1.44-2.85) | <.001 | 1.94 (1.38-2.73) | <.001 | 1.80 (1.31-2.46) | <.001 | 1.87 (1.33-2.64) | <.001 | 1.78 (1.26-2.51) | .001 | 1.41 (1.04-1.90) | .028 | 1.51 (1.09-2.11) | .015 | 1.45 (1.04-2.03) | .029 |
| 47 | 2.26 (1.57-3.26) | <.001 | 2.09 (1.44-3.04) | <.001 | 2.00 (1.37-2.91) | <.001 | 2.13 (1.52-2.98) | <.001 | 1.88 (1.29-2.74) | .001 | 1.75 (1.19-2.55) | .004 | 1.58 (1.14-2.19) | .006 | 1.28 (0.89-1.85) | .182 | 1.19 (0.82-1.73) | .357 |
| 48 | 3.06 (2.05-4.55) | <.001 | 2.83 (1.88-4.26) | <.001 | 2.72 (1.80-4.11) | <.001 | 1.69 (1.17-2.45) | .006 | 2.03 (1.34-3.09) | <.001 | 1.91 (1.26-2.91) | .002 | 1.47 (1.03-2.11) | .034 | 1.35 (0.90-2.02) | .144 | 1.27 (0.85-1.90) | .250 |
| 49 | 3.54 (2.26-5.57) | <.001 | 3.27 (2.06-5.19) | <.001 | 3.02 (1.90-4.82) | <.001 | 1.92 (1.28-2.88) | .002 | 2.30 (1.45-3.67) | <.001 | 2.16 (1.35-3.45) | .001 | 1.49 (1.01-2.20) | .047 | 1.25 (0.79-1.96) | .338 | 1.16 (0.73-1.84) | .519 |
| 50 | 3.65 (2.23-5.95) | <.001 | 3.46 (2.10-5.72) | <.001 | 3.09 (1.86-5.14) | <.001 | 2.06 (1.30-3.25) | .002 | 2.04 (1.23-3.40) | .006 | 1.82 (1.08-3.06) | .023 | 1.24 (0.80-1.93) | .339 | 1.12 (0.69-1.84) | .647 | 1.01 (0.61-1.68) | .956 |
| 51 | 4.29 (2.49-7.39) | <.001 | 4.22 (2.42-7.34) | <.001 | 3.89 (2.24-6.77) | <.001 | 1.96 (1.20-3.22) | .008 | 2.16 (1.25-3.73) | .006 | 2.00 (1.16-3.46) | .013 | 1.27 (0.79-2.06) | .322 | 1.31 (0.76-2.25) | .333 | 1.28 (0.74-2.20) | .380 |
| 52 | 4.25 (2.27-7.96) | <.001 | 4.09 (2.14-7.82) | <.001 | 3.67 (1.92-7.03) | <.001 | 2.58 (1.52-4.37) | <.001 | 2.72 (1.44-5.11) | .002 | 2.48 (1.32-4.68) | .005 | 1.37 (0.81-2.33) | .238 | 1.22 (0.64-2.32) | .546 | 1.19 (0.62-2.27) | .604 |
| 53 | 6.93 (3.31-14.51) | <.001 | 5.82 (2.68-12.65) | <.001 | 5.82 (2.68-12.65) | <.001 | 3.05 (1.66-5.62) | <.001 | 3.54 (1.71-7.33) | <.001 | 3.54 (1.71-7.33) | <.001 | 1.56 (0.85-2.86) | .151 | 1.76 (0.84-3.68) | .134 | 1.76 (0.84-3.68) | .134 |
| 54 | 9.36 (3.97-22.05) | <.001 | 7.71 (3.21-18.50) | <.001 | 7.71 (3.21-18.50) | <.001 | 4.68 (2.34-9.36) | <.001 | 4.02 (1.81-8.92) | <.001 | 4.02 (1.81-8.92) | <.001 | 1.79 (0.90-3.58) | .099 | 1.47 (0.67-3.20) | .337 | 1.47 (0.67-3.20) | .337 |
| 55 | 14.49 (5.29-39.71) | <.001 | 12.61 (4.53-35.06) | <.001 | 12.61 (4.53-35.06) | <.001 | 5.05 (2.34-10.89) | <.001 | 4.79 (2.00-11.45) | <.001 | 4.79 (2.00-11.45) | <.001 | 1.96 (0.92-4.20) | .083 | 1.88 (0.79-4.46) | .155 | 1.88 (0.79-4.46) | .155 |

Model 1 (crude) adjusts for onset age, sex, birth cohort, parental psychiatric history, and parental socioeconomic status. Model 2 additionally adjusts for active anorexia nervosa diagnostic status at the corresponding year. Model 3 additionally adjusts for active eating disorder diagnostic status at the corresponding year.

**eTable 5.** Age-Specific Odds Ratios for Employment, Income, and Cohabitation Outcomes by Anorexia Nervosa Severity Quintile, Relative to the General Population

| **Age** | **Work** | | | **Income** | | **Cohabitation** | |
| --- | --- | --- | --- | --- | --- | --- | --- |
|  | **Group** | **OR (95% CI)** | **P value** | **OR (95% CI)** | **P value** | **OR (95% CI)** | **P value** |
| 30 | GP | 1 |  | 1 |  | 1 |  |
|  | Quintile 1 | 1.59 (1.39-1.83) | <.001 | 1.97 (1.74-2.24) | <.001 | 1.59 (1.43-1.78) | <.001 |
|  | Quintile 2 | 1.35 (1.10-1.65) | .003 | 1.41 (1.16-1.72) | <.001 | 1.37 (1.17-1.59) | <.001 |
|  | Quintile 3 | 1.88 (1.66-2.12) | <.001 | 1.91 (1.69-2.15) | <.001 | 1.54 (1.38-1.71) | <.001 |
|  | Quintile 4 | 2.29 (2.01-2.61) | <.001 | 2.28 (2.00-2.59) | <.001 | 1.83 (1.63-2.05) | <.001 |
|  | Quintile 5 | 3.42 (3.03-3.86) | <.001 | 3.84 (3.42-4.31) | <.001 | 2.43 (2.17-2.71) | <.001 |
| 31 | GP | 1 |  | 1 |  | 1 |  |
|  | Quintile 1 | 1.66 (1.44-1.92) | <.001 | 1.85 (1.61-2.12) | <.001 | 1.62 (1.44-1.82) | <.001 |
|  | Quintile 2 | 1.55 (1.26-1.90) | <.001 | 1.66 (1.36-2.02) | <.001 | 1.37 (1.16-1.62) | <.001 |
|  | Quintile 3 | 2.02 (1.78-2.28) | <.001 | 2.08 (1.84-2.35) | <.001 | 1.52 (1.36-1.70) | <.001 |
|  | Quintile 4 | 2.26 (1.97-2.59) | <.001 | 2.41 (2.11-2.76) | <.001 | 1.85 (1.64-2.09) | <.001 |
|  | Quintile 5 | 3.52 (3.10-3.99) | <.001 | 3.68 (3.26-4.15) | <.001 | 2.42 (2.16-2.72) | <.001 |
| 32 | GP | 1 |  | 1 |  | 1 |  |
|  | Quintile 1 | 1.90 (1.64-2.20) | <.001 | 1.96 (1.70-2.26) | <.001 | 1.68 (1.48-1.90) | <.001 |
|  | Quintile 2 | 1.56 (1.25-1.93) | <.001 | 1.61 (1.30-1.99) | <.001 | 1.35 (1.13-1.61) | .001 |
|  | Quintile 3 | 2.02 (1.78-2.30) | <.001 | 2.05 (1.81-2.33) | <.001 | 1.51 (1.35-1.69) | <.001 |
|  | Quintile 4 | 2.38 (2.07-2.74) | <.001 | 2.34 (2.04-2.69) | <.001 | 1.81 (1.60-2.06) | <.001 |
|  | Quintile 5 | 3.72 (3.26-4.25) | <.001 | 3.61 (3.18-4.10) | <.001 | 2.49 (2.21-2.81) | <.001 |
| 33 | GP | 1 |  | 1 |  | 1 |  |
|  | Quintile 1 | 2.09 (1.80-2.44) | <.001 | 1.89 (1.63-2.20) | <.001 | 1.79 (1.57-2.04) | <.001 |
|  | Quintile 2 | 1.67 (1.33-2.09) | <.001 | 1.65 (1.32-2.06) | <.001 | 1.50 (1.24-1.81) | <.001 |
|  | Quintile 3 | 2.24 (1.96-2.55) | <.001 | 2.10 (1.84-2.40) | <.001 | 1.55 (1.38-1.75) | <.001 |
|  | Quintile 4 | 2.60 (2.25-3.01) | <.001 | 2.41 (2.08-2.79) | <.001 | 1.88 (1.65-2.15) | <.001 |
|  | Quintile 5 | 4.08 (3.56-4.68) | <.001 | 3.60 (3.14-4.11) | <.001 | 2.43 (2.14-2.76) | <.001 |
| 34 | GP | 1 |  | 1 |  | 1 |  |
|  | Quintile 1 | 2.01 (1.70-2.37) | <.001 | 1.88 (1.60-2.21) | <.001 | 1.71 (1.49-1.97) | <.001 |
|  | Quintile 2 | 1.53 (1.20-1.96) | <.001 | 1.75 (1.38-2.21) | <.001 | 1.43 (1.17-1.75) | <.001 |
|  | Quintile 3 | 2.32 (2.03-2.66) | <.001 | 2.25 (1.96-2.57) | <.001 | 1.60 (1.41-1.81) | <.001 |
|  | Quintile 4 | 2.77 (2.38-3.22) | <.001 | 2.56 (2.21-2.98) | <.001 | 2.07 (1.81-2.38) | <.001 |
|  | Quintile 5 | 4.09 (3.54-4.73) | <.001 | 3.52 (3.06-4.05) | <.001 | 2.33 (2.04-2.66) | <.001 |
| 35 | GP | 1 |  | 1 |  | 1 |  |
|  | Quintile 1 | 2.19 (1.84-2.59) | <.001 | 1.93 (1.63-2.27) | <.001 | 1.78 (1.54-2.07) | <.001 |
|  | Quintile 2 | 1.91 (1.50-2.44) | <.001 | 1.60 (1.25-2.04) | <.001 | 1.37 (1.10-1.69) | .004 |
|  | Quintile 3 | 2.26 (1.96-2.60) | <.001 | 2.09 (1.82-2.40) | <.001 | 1.72 (1.51-1.95) | <.001 |
|  | Quintile 4 | 3.14 (2.68-3.67) | <.001 | 2.60 (2.24-3.03) | <.001 | 2.04 (1.76-2.35) | <.001 |
|  | Quintile 5 | 4.01 (3.45-4.67) | <.001 | 3.14 (2.70-3.64) | <.001 | 2.29 (1.98-2.63) | <.001 |
| 36 | GP | 1 |  | 1 |  | 1 |  |
|  | Quintile 1 | 2.31 (1.94-2.77) | <.001 | 2.25 (1.90-2.67) | <.001 | 1.78 (1.52-2.08) | <.001 |
|  | Quintile 2 | 2.03 (1.58-2.62) | <.001 | 1.67 (1.30-2.16) | <.001 | 1.34 (1.06-1.67) | .011 |
|  | Quintile 3 | 2.11 (1.82-2.45) | <.001 | 2.03 (1.75-2.34) | <.001 | 1.70 (1.49-1.93) | <.001 |
|  | Quintile 4 | 3.17 (2.70-3.74) | <.001 | 2.68 (2.28-3.15) | <.001 | 2.13 (1.83-2.47) | <.001 |
|  | Quintile 5 | 4.30 (3.67-5.04) | <.001 | 3.32 (2.85-3.88) | <.001 | 2.20 (1.90-2.56) | <.001 |
| 37 | GP | 1 |  | 1 |  | 1 |  |
|  | Quintile 1 | 2.29 (1.90-2.76) | <.001 | 1.96 (1.63-2.35) | <.001 | 1.85 (1.57-2.17) | <.001 |
|  | Quintile 2 | 1.68 (1.27-2.22) | <.001 | 1.63 (1.24-2.13) | <.001 | 1.27 (1.00-1.61) | .048 |
|  | Quintile 3 | 2.19 (1.87-2.56) | <.001 | 2.04 (1.75-2.37) | <.001 | 1.68 (1.47-1.93) | <.001 |
|  | Quintile 4 | 3.05 (2.57-3.63) | <.001 | 2.67 (2.26-3.16) | <.001 | 2.02 (1.72-2.36) | <.001 |
|  | Quintile 5 | 4.45 (3.77-5.26) | <.001 | 3.26 (2.77-3.84) | <.001 | 2.24 (1.91-2.61) | <.001 |
| 38 | GP | 1 |  | 1 |  | 1 |  |
|  | Quintile 1 | 2.22 (1.82-2.71) | <.001 | 2.04 (1.68-2.46) | <.001 | 1.75 (1.47-2.07) | <.001 |
|  | Quintile 2 | 1.90 (1.42-2.53) | <.001 | 1.88 (1.44-2.46) | <.001 | 1.33 (1.04-1.70) | .023 |
|  | Quintile 3 | 2.22 (1.89-2.61) | <.001 | 2.00 (1.71-2.34) | <.001 | 1.61 (1.40-1.86) | <.001 |
|  | Quintile 4 | 3.24 (2.70-3.90) | <.001 | 2.60 (2.18-3.10) | <.001 | 2.08 (1.76-2.45) | <.001 |
|  | Quintile 5 | 4.29 (3.60-5.11) | <.001 | 3.15 (2.65-3.74) | <.001 | 2.29 (1.95-2.70) | <.001 |
| 39 | GP | 1 |  | 1 |  | 1 |  |
|  | Quintile 1 | 2.37 (1.92-2.92) | <.001 | 2.01 (1.64-2.46) | <.001 | 1.76 (1.47-2.11) | <.001 |
|  | Quintile 2 | 1.92 (1.40-2.62) | <.001 | 1.90 (1.43-2.52) | <.001 | 1.33 (1.02-1.72) | .031 |
|  | Quintile 3 | 2.27 (1.92-2.68) | <.001 | 2.14 (1.82-2.51) | <.001 | 1.71 (1.48-1.98) | <.001 |
|  | Quintile 4 | 3.18 (2.62-3.85) | <.001 | 2.65 (2.20-3.20) | <.001 | 2.02 (1.70-2.41) | <.001 |
|  | Quintile 5 | 4.74 (3.94-5.71) | <.001 | 3.40 (2.84-4.06) | <.001 | 2.23 (1.88-2.65) | <.001 |
| 40 | GP | 1 |  | 1 |  | 1 |  |
|  | Quintile 1 | 2.63 (2.11-3.27) | <.001 | 2.14 (1.73-2.63) | <.001 | 2.06 (1.70-2.49) | <.001 |
|  | Quintile 2 | 2.40 (1.74-3.30) | <.001 | 2.06 (1.53-2.78) | <.001 | 1.20 (0.89-1.59) | .222 |
|  | Quintile 3 | 2.12 (1.77-2.52) | <.001 | 1.98 (1.67-2.34) | <.001 | 1.74 (1.50-2.02) | <.001 |
|  | Quintile 4 | 3.32 (2.72-4.06) | <.001 | 2.81 (2.33-3.41) | <.001 | 1.99 (1.66-2.39) | <.001 |
|  | Quintile 5 | 4.98 (4.10-6.04) | <.001 | 3.83 (3.18-4.62) | <.001 | 2.44 (2.03-2.93) | <.001 |
| 41 | GP | 1 |  | 1 |  | 1 |  |
|  | Quintile 1 | 2.60 (2.06-3.29) | <.001 | 2.18 (1.75-2.72) | <.001 | 2.10 (1.72-2.57) | <.001 |
|  | Quintile 2 | 2.31 (1.64-3.25) | <.001 | 2.11 (1.53-2.90) | <.001 | 1.27 (0.93-1.72) | .126 |
|  | Quintile 3 | 2.21 (1.85-2.65) | <.001 | 1.90 (1.59-2.26) | <.001 | 1.82 (1.56-2.12) | <.001 |
|  | Quintile 4 | 3.38 (2.74-4.18) | <.001 | 2.99 (2.45-3.64) | <.001 | 1.99 (1.64-2.41) | <.001 |
|  | Quintile 5 | 4.83 (3.93-5.93) | <.001 | 3.83 (3.15-4.65) | <.001 | 2.39 (1.97-2.89) | <.001 |
| 42 | GP | 1 |  | 1 |  | 1 |  |
|  | Quintile 1 | 2.79 (2.17-3.58) | <.001 | 2.25 (1.78-2.86) | <.001 | 2.21 (1.78-2.74) | <.001 |
|  | Quintile 2 | 2.44 (1.69-3.54) | <.001 | 2.03 (1.44-2.86) | <.001 | 1.09 (0.77-1.51) | .623 |
|  | Quintile 3 | 2.13 (1.76-2.57) | <.001 | 2.03 (1.69-2.42) | <.001 | 1.74 (1.48-2.05) | <.001 |
|  | Quintile 4 | 3.18 (2.54-3.97) | <.001 | 2.67 (2.16-3.30) | <.001 | 2.05 (1.67-2.49) | <.001 |
|  | Quintile 5 | 4.57 (3.66-5.70) | <.001 | 3.62 (2.94-4.46) | <.001 | 2.34 (1.91-2.86) | <.001 |
| 43 | GP | 1 |  | 1 |  | 1 |  |
|  | Quintile 1 | 3.11 (2.39-4.07) | <.001 | 2.66 (2.08-3.40) | <.001 | 2.18 (1.73-2.73) | <.001 |
|  | Quintile 2 | 2.46 (1.64-3.69) | <.001 | 2.16 (1.49-3.12) | <.001 | 1.52 (1.07-2.13) | .017 |
|  | Quintile 3 | 2.10 (1.72-2.56) | <.001 | 1.79 (1.48-2.17) | <.001 | 1.85 (1.57-2.18) | <.001 |
|  | Quintile 4 | 3.09 (2.44-3.92) | <.001 | 2.72 (2.18-3.40) | <.001 | 2.05 (1.66-2.52) | <.001 |
|  | Quintile 5 | 4.53 (3.56-5.76) | <.001 | 3.39 (2.70-4.25) | <.001 | 2.28 (1.83-2.83) | <.001 |
| 44 | GP | 1 |  | 1 |  | 1 |  |
|  | Quintile 1 | 3.04 (2.25-4.10) | <.001 | 2.13 (1.61-2.81) | <.001 | 2.05 (1.59-2.62) | <.001 |
|  | Quintile 2 | 2.57 (1.65-4.00) | <.001 | 2.13 (1.42-3.20) | <.001 | 1.26 (0.84-1.83) | .247 |
|  | Quintile 3 | 2.27 (1.86-2.77) | <.001 | 1.94 (1.60-2.36) | <.001 | 1.83 (1.54-2.17) | <.001 |
|  | Quintile 4 | 3.19 (2.49-4.08) | <.001 | 2.36 (1.86-3.01) | <.001 | 2.01 (1.61-2.50) | <.001 |
|  | Quintile 5 | 4.85 (3.76-6.25) | <.001 | 3.48 (2.73-4.44) | <.001 | 2.46 (1.95-3.11) | <.001 |
| 45 | GP | 1 |  | 1 |  | 1 |  |
|  | Quintile 1 | 2.66 (1.87-3.80) | <.001 | 2.13 (1.55-2.92) | <.001 | 2.13 (1.61-2.81) | <.001 |
|  | Quintile 2 | 2.88 (1.78-4.65) | <.001 | 2.32 (1.48-3.62) | <.001 | 1.23 (0.79-1.87) | .340 |
|  | Quintile 3 | 2.16 (1.75-2.66) | <.001 | 2.03 (1.66-2.48) | <.001 | 1.75 (1.46-2.09) | <.001 |
|  | Quintile 4 | 2.94 (2.26-3.82) | <.001 | 2.45 (1.90-3.15) | <.001 | 2.03 (1.61-2.56) | <.001 |
|  | Quintile 5 | 4.65 (3.54-6.10) | <.001 | 3.98 (3.08-5.14) | <.001 | 2.50 (1.94-3.21) | <.001 |
| 46 | GP | 1 |  | 1 |  | 1 |  |
|  | Quintile 1 | 2.15 (1.40-3.30) | <.001 | 1.86 (1.28-2.72) | .001 | 1.85 (1.33-2.55) | <.001 |
|  | Quintile 2 | 2.51 (1.46-4.31) | <.001 | 2.57 (1.58-4.16) | <.001 | 1.53 (0.95-2.41) | .071 |
|  | Quintile 3 | 2.25 (1.81-2.80) | <.001 | 2.00 (1.62-2.47) | <.001 | 1.72 (1.42-2.07) | <.001 |
|  | Quintile 4 | 3.12 (2.37-4.10) | <.001 | 2.36 (1.80-3.09) | <.001 | 2.17 (1.70-2.77) | <.001 |
|  | Quintile 5 | 5.20 (3.89-6.96) | <.001 | 3.75 (2.85-4.93) | <.001 | 2.57 (1.96-3.36) | <.001 |
| 47 | GP | 1 |  | 1 |  | 1 |  |
|  | Quintile 1 | 1.77 (1.08-2.91) | .025 | 1.60 (1.02-2.53) | .042 | 1.62 (1.10-2.35) | .013 |
|  | Quintile 2 | 2.56 (1.42-4.63) | .002 | 2.57 (1.52-4.37) | <.001 | 1.62 (0.97-2.67) | .062 |
|  | Quintile 3 | 2.11 (1.68-2.65) | <.001 | 1.94 (1.55-2.42) | <.001 | 1.62 (1.33-1.97) | <.001 |
|  | Quintile 4 | 3.11 (2.32-4.18) | <.001 | 2.62 (1.98-3.46) | <.001 | 2.24 (1.72-2.90) | <.001 |
|  | Quintile 5 | 5.27 (3.82-7.26) | <.001 | 4.38 (3.27-5.86) | <.001 | 2.79 (2.09-3.74) | <.001 |
| 48 | GP | 1 |  | 1 |  | 1 |  |
|  | Quintile 1 | 2.31 (1.33-4.03) | .003 | 1.44 (0.86-2.43) | .170 | 1.28 (0.82-1.96) | .273 |
|  | Quintile 2 | 1.98 (0.96-4.11) | .066 | 2.74 (1.53-4.91) | <.001 | 1.43 (0.79-2.49) | .222 |
|  | Quintile 3 | 1.98 (1.56-2.53) | <.001 | 2.07 (1.65-2.60) | <.001 | 1.57 (1.27-1.92) | <.001 |
|  | Quintile 4 | 2.65 (1.92-3.65) | <.001 | 2.52 (1.87-3.40) | <.001 | 2.37 (1.79-3.12) | <.001 |
|  | Quintile 5 | 6.48 (4.57-9.18) | <.001 | 3.55 (2.56-4.92) | <.001 | 2.54 (1.85-3.50) | <.001 |
| 49 | GP | 1 |  | 1 |  | 1 |  |
|  | Quintile 1 | 2.48 (1.20-5.12) | .014 | 1.60 (0.88-2.92) | .123 | 1.77 (1.07-2.90) | .025 |
|  | Quintile 2 | 1.80 (0.76-4.29) | .184 | 1.67 (0.79-3.55) | .178 | 1.59 (0.81-3.04) | .170 |
|  | Quintile 3 | 1.99 (1.54-2.56) | <.001 | 1.99 (1.57-2.53) | <.001 | 1.53 (1.23-1.90) | <.001 |
|  | Quintile 4 | 3.04 (2.17-4.26) | <.001 | 2.58 (1.88-3.53) | <.001 | 2.43 (1.81-3.28) | <.001 |
|  | Quintile 5 | 7.78 (5.22-11.60) | <.001 | 3.80 (2.67-5.41) | <.001 | 2.68 (1.89-3.81) | <.001 |
| 50 | GP | 1 |  | 1 |  | 1 |  |
|  | Quintile 1 | 1.45 (0.54-3.90) | .461 | 1.20 (0.50-2.91) | .684 | 1.47 (0.73-2.84) | .263 |
|  | Quintile 2 | 2.80 (1.02-7.69) | .045 | 1.96 (0.83-4.63) | .127 | 1.78 (0.81-3.79) | .140 |
|  | Quintile 3 | 1.83 (1.40-2.40) | <.001 | 2.06 (1.60-2.65) | <.001 | 1.62 (1.29-2.03) | <.001 |
|  | Quintile 4 | 3.11 (2.16-4.48) | <.001 | 3.11 (2.22-4.36) | <.001 | 2.76 (2.00-3.83) | <.001 |
|  | Quintile 5 | 7.50 (4.87-11.54) | <.001 | 4.66 (3.13-6.92) | <.001 | 2.39 (1.61-3.55) | <.001 |

GP indicates general population; quintiles are derived from the AN Register-based Severity Index (Q1, lowest, to Q5, highest).
